# The effect of intimate partner violence on women’s risk of HIV acquisition and engagement in the HIV treatment and care cascade: an individual-participant data meta-analysis of nationally representative surveys in sub-Saharan Africa

**DOI:** 10.1101/2022.08.04.22278331

**Authors:** S Kuchukhidze, D Panagiotoglou, MC Boily, S Diabaté, JW Eaton, F Mbofana, L Sardinha, L Schrubbe, H Stöckl, RK Wanyenze, M Maheu-Giroux

**Author notes:** **Corresponding author** Mathieu Maheu-Giroux, 2001 Avenue McGill College, Suite 1200, Montréal, Québec, H3A 1G1, CANADA.

## Abstract

**Background:** Achieving the 95-95-95 targets for HIV diagnosis, treatment, and viral load suppression (VLS) to end the HIV/AIDS epidemic hinges on eliminating structural inequalities, including intimate partner violence (IPV). Sub-Saharan Africa (SSA) has among the world’s highest prevalence of IPV and HIV. We aim to examine the impact of IPV on recent HIV infection and women’s engagement in the HIV care cascade.

**Methods:** We pooled individual-level data from nationally representative surveys with information on physical and/or sexual IPV and HIV in SSA (2000-2020). We used Poisson regressions with robust standard errors to estimate adjusted prevalence ratios (aPR) of past year experience of IPV on recent HIV infection (measured using recency assays), past-year HIV testing, antiretroviral therapy (ART) uptake, and VLS among ever-partnered (currently/formerly married or cohabitating) women. Models were adjusted for women’s age, age at sexual debut, residence type, marital status, education, and the survey’s identifier (proxy of country and year).

**Findings:** Fifty-seven surveys were available from thirty countries, encompassing 280,259 (N_i_) individuals. One-fifth of women reported physical and/or sexual IPV in the past year. Six surveys had information on recent HIV infection and seven had data on ART uptake and VLS. Women experiencing past year IPV were 3·22 times (95%CI: 1·51-6·85, N_i_=19,179) more likely to have a recent HIV infection, adjusting for potential confounders. Past year IPV was not associated with HIV testing (aPR=0·99, 95%CI: 0·98-1·01, N_i_=274,506) or ART uptake (aPR=0·96, 95%CI:0·90-1·03, N_i_=5,629). Women living with HIV (WLHIV) experiencing IPV in the past year were 9% less likely to achieve VLS (aPR=0·91, 95%CI: 0·85-0·98, N_i_=5,627).

**Interpretation:** Past year IPV was associated with recent HIV acquisition and lower VLS among WLHIV. Preventing IPV is inherently imperative but eliminating IPV could also be crucial to ending HIV/AIDS.

**Funding:** Canadian Institutes of Health Research, Canada Research Chair, and Max E. Binz Fellowship

Research in Context

Evidence before this study
Our study builds on more than two decades of research devoted to IPV and HIV.^1-4^ We summarized this scholarship by searching PubMed for empirical studies (April 8, 2022), without language restrictions using the terms: HIV AND women AND (violence OR intimate partner OR domestic violence OR GBV OR IPV) AND (Africa* OR sub-Sahara*).
Several systematic and scoping reviews have been conducted on the links between IPV and HIV with mixed results.^5-7^ Most studies used HIV seropositivity as the outcome. In sub-Saharan Africa, a multi-country study found no association between IPV and HIV; however it has been subsequently suggested that women experiencing IPV have an increased risk of HIV seropositivity depending on the chosen referent group.^8^ Longitudinal studies in Uganda and South Africa suggest that women experiencing IPV are more likely to acquire HIV compared to those who are not.^9,10^ However, two other cohort studies among youth^11^ and serodiscordant couples^12^ did not find significant associations.
Impacts of IPV on the HIV prevention and treatment cascade have also been studied. A 2015 meta-analysis of 13 cross-sectional studies, mostly from the United States, found that IPV is associated with lower ART use, adherence and VLS.^13^ For the first step in the cascade – HIV diagnosis– most included studies in a 2019 scoping review by Leddy^14^ did not find an association between IPV and HIV testing^15-19^; though two reported a reduction in testing associated with IPV among pregnant and postpartum women.^20,21^
Evidence supports a negative relationship between IPV and ART adherence, though most studies focused on populations such as sex workers^17^, youth^22^, or pregnant/postpartum women.^18,23-25^ Quantitative evidence are sparse on ART *uptake*, but among five studies from high income settings, none showed an association between IPV and *current* ART use.^13^ Studies from Zambia and South Africa point to an association between IPV and unsuppressed viral loads among adolescents/youth^26^ and postpartum women.^27^
Overall, comparison of estimates and outcomes is difficult due to a lack of standardization in survey instruments, recall period for IPV, considered IPV types, outcome measurement, and populations considered (e.g., pregnant and/or young women, sex workers).

Added value of this study
Using microdata from population-based surveys in the region with the highest HIV burden, we conducted the largest and most comprehensive study to date on the impacts of IPV on HIV: from HIV acquisition to the three most important steps of the treatment and care cascade. Our results generally corroborate previous findings, but we considerably expanded the scope of previous studies using recent experiences of IPV, consideration of partner characteristics, and estimating their impact on biomarker-based measures of HIV acquisition, ART coverage, and VLS.

The implications of all the available evidence
The *2021 Political Declaration on AIDS* commits to eliminating sexual and gender-based violence, including IPV as key to combatting the AIDS epidemic. IPV is an urgent public health concern, and a barrier to the elimination of AIDS. Our results highlight the need for HIV programs to consider IPV along the full continuum of HIV prevention to treatment and care. Both IPV and HIV are preventable. Governments, societies, and communities need to explicitly recognize the IPV-HIV syndemic, and implement multisectoral response with social and economic components to eliminate gender-based violence and end AIDS.

## Introduction

Despite significant progress to curb HIV epidemics worldwide, 1.5 million new HIV infections occurred in 2020, over 60% of which were in sub-Saharan Africa.^28^ This burden of new infections disproportionately affects women: they account for 65% of new HIV infections in sub-Saharan Africa.^28,29^ The global HIV agenda is guided by the “95-95-95” targets to end AIDS by 2030 — an ambitious plan that calls for achieving 95% diagnosis coverage, 95% antiretroviral therapy (ART) uptake among those diagnosed, and 95% of viral suppression among those on treatement.^30^ Reaching these targets partly hinges upon addressing structural vulnerabilities such as inequitable gender and social norms, and violence against women and girls. Worldwide, over one in four women has experienced physical and/or sexual intimate partner violence (IPV) in their lifetime, with prevalence reaching highs of approximately 40% in Central and Eastern sub-Saharan Africa.^31^ This violence often co-occurs with HIV and could function as a syndemic, posing barriers to women’s ability to prevent HIV acquisition, to access HIV care, and to remain in care if living with the virus. The *2021 United Nations General Assembly* adopted the *Political Declaration on HIV and AIDS* with bold new global targets for 2025, which commit to elimination of all forms of sexual and gender-based violence, including IPV as a key enabler of the HIV epidemic.^32^ Improving our understanding of the complex relationships between IPV and HIV is essential to meet this commitment.

In sub-Saharan Africa, women being subjected to IPV could be at increased risk of HIV acquisition.^8-10^ Beside this potential impact on HIV incidence, IPV could compromise access to the HIV prevention and care cascade^33^: from HIV testing, to ART uptake and retention^14,17,22,34^, and ultimately, to viral load suppression (VLS).^13,14,26^ Overall, evidence suggesting that IPV and HIV interact in a syndemic way could be strengthened. Previous studies either provided mixed results, focused on a single country, and/or recruited specific populations such as pregnant women, ^34^ youth^22^, female sex workers^17^, or women who use substances.^35^ This makes generalization of the study’s results challenging. Furthermore, the definitions of IPV (e.g., severity of acts, physical only, sexual only, or both), the period (e.g., lifetime or past year), and sampling strategies (e.g., currently partnered or ever-partnered) have varied, making it difficult to systematically compare effect size estimates, and generate robust evidence on population-level effects of IPV.^14^ Over the last decade, several large, nationally-representative, population-based surveys have collected information on IPV and HIV, including recency assays, ART biomarkers, and VLS. These surveys use standardized and robust methodology, providing researchers opportunities to overcome the limitations of previous studies.

Our overarching aim was to improve understanding of the complex relationships between women’s experience of IPV and HIV acquisition, and engagement with the HIV prevention and treatment cascade.^2,3,33^ To achieve this, we estimated the impact of recent (past 12 months) physical and/or sexual IPV on the following four outcomes: recent HIV infection, self-reported HIV testing in the past year, ART uptake, and VLS.

## Methods

### Data sources and study population

We reviewed all available nationally-representative, cross-sectional, population-based surveys from sub-Saharan Africa over 2000-2020 with individual participant data on both IPV and HIV. We searched data catalogs (i.e., the *Global Health Data Exchange*, the *International Household Survey Network*), examined surveys included in the Global Estimates for Violence Against Women Statistics systematic review^36^, a previous review of surveys with information on HIV testing^37^, and complemented these with expert knowledge.

The types of surveys considered included *Demographic and Health Surveys* (DHS), *AIDS Indicator Surveys* (AIS) (https://dhsprogram.com/Data/), *Population-based HIV Impact Assessment* (PHIA) (https://phia-data.icap.columbia.edu/datasets) and *South Africa National HIV Prevalence, Incidence, Behavior and Communication Survey* (SABSSM) (http://datacuration.hsrc.ac.za/search/browse/alpha/S), as well as country-specific surveys (e.g., *Botswana AIDS Impact Survey, Kenya AIDS Indicator Surveys*, and *Nigeria HIV/AIDS Indicator and Impact Survey*). Study population included all ever-partnered (currently or formerly married or cohabitating) women and girls aged ≥15 years (Figure 1).

**Figure 1.**
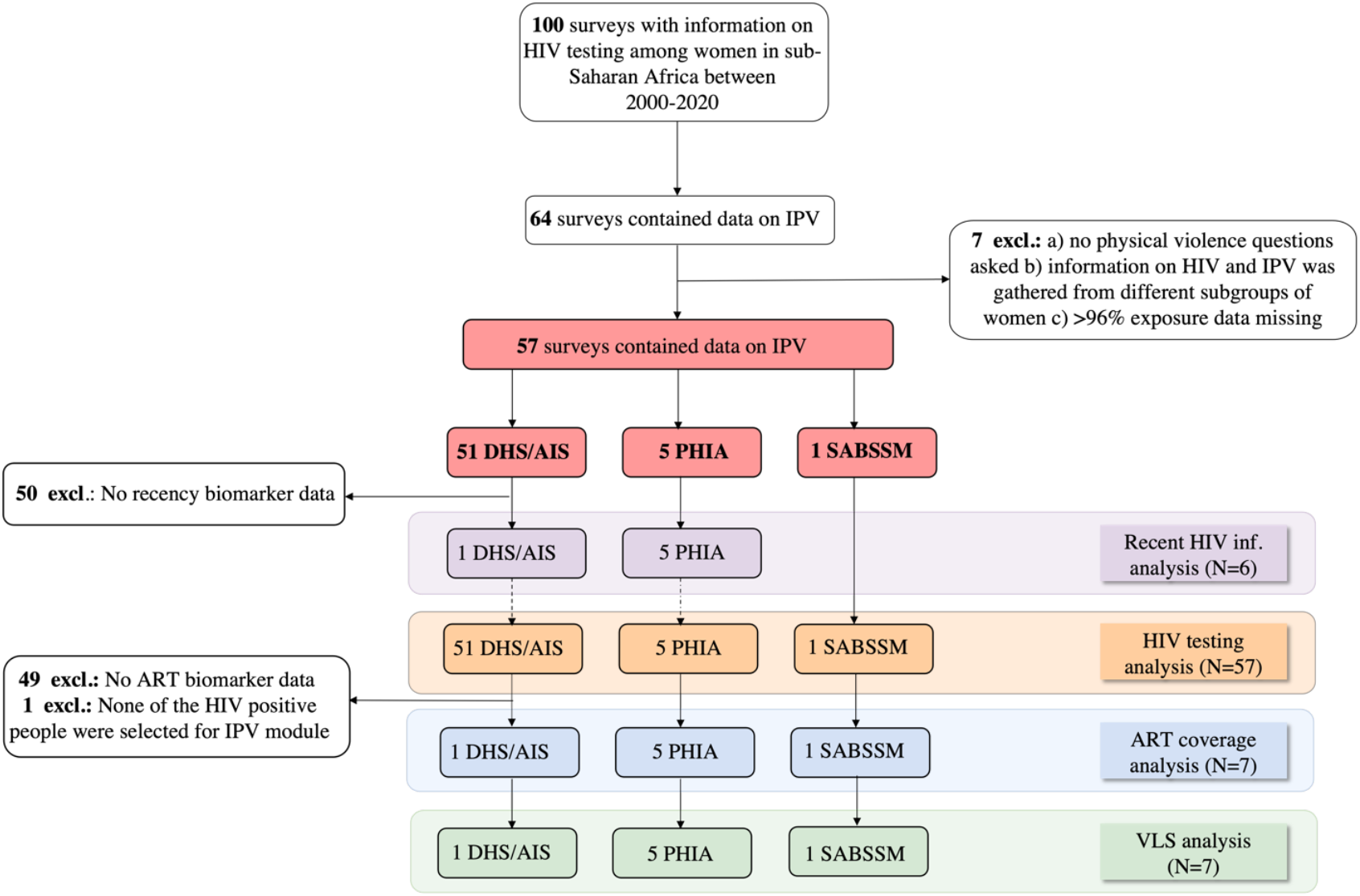
Flowchart for survey inclusion/exclusion for each type of analysis: recent HIV infection, HIV testing, antiretroviral (ART) uptake and viral load suppression (VLS). *We did not use 7 surveys with data on intimate partner violence (IPV) since information on past year physical IPV were not collected (N*_*surv*_*=3) or information about HIV and intimate partner violence was gathered from different subgroups of women (N*_*surv*_*=2) (i*.*e*., *HIV questions in the DHS were asked to women in households which were selected for the men’s survey). We removed two surveys (Côte d’Ivoire PHIA 2017 and Cameroon PHIA 2017) where over 95% of past year IPV data were missing*. (AIS=AIDS Indicator Survey; DHS=Demographic and Health Survey; Excl.=excluded; IPV=intimate partner violence; PHIA=Population-based HIV Impact Assessment Survey; SABSSM=South African National HIV Prevalence, Incidence, Behaviour and Communication Survey.)

### Measurement of experience of IPV

In all surveys, data on IPV was collected from one randomly selected woman in each household for PHIA and SABSSM, and from a fraction of households in DHS/AIS (usually one third). The primary exposure of interest was experience of physical and/or sexual IPV in the past year (Supplement 1; Table S1). All surveys used acts-specific, gold-standard instruments based on the modified *Conflict Tactics Scale* to collect information on IPV.^38,39^

The secondary exposures of interest were a) lifetime experience of physical IPV only, b) lifetime experience of sexual IPV only, c) lifetime experience of physical and/or sexual IPV, d) lifetime experience of severe physical and/or sexual IPV, and d) frequent past year experience of physical and/or sexual IPV. Measurements are generally consistent across surveys, although PHIAs collected information on past year IPV only (no lifetime measure) (Supplement 1; Table S1) and in SABSSM, past year IPV pertains to physical violence only (i.e., no information on sexual violence).

Severity of lifetime IPV was collected only in the DHS and SABSSM (number of surveys, N_surv_=52). Severe IPV was defined as lifetime experience of severe physical and/or sexual IPV, which includes punching, kicking/dragging, trying to strangle/burn, threatening with a weapon, attacking with a weapon, and any type of sexual IPV.^40^ (Supplement 1; Table S1)

Frequency of past year physical and/or sexual IPV was categorized as “often”, “sometimes”, or “never”. In PHIA, the frequency question pertains to the experience of physical violence by any perpetrator in the past year, and we assumed this to be an intimate partner only if the participant reported IPV as well (Supplement 1; Table S1). Further, in the SABSSM and PHIA surveys, the frequency of past year IPV pertains to only physical IPV while in other surveys the indicator refers to frequency of physical and/or sexual IPV. Whenever the survey did not collect the information, we extrapolated the frequency of physical IPV to that of physical and/or sexual IPV based on the strong relationship between both measures (X-squared = 43,780, p<0.05).

### Outcome measurements

Our primary outcome is recent HIV infection (as a proxy for HIV incidence) among women at risk of HIV acquisition (i.e., excluding those living with non-recent HIV). Recency was measured via LAg assay performed on all participants found to be seropositive for HIV. The algorithm to identify recent infections –those that were acquired less than four to seven months before sample collection^41,42^– accounted for ART biomarkers and VLS to minimize false positives.^43^

Other outcomes concern the HIV prevention and treatment cascade. (Supplement 1; Table S2) First, we considered self-reported HIV testing histories (lifetime and past year testing and receipt of result). Second, ART uptake was measured among ever-partnered women living with HIV (WLHIV) based on either ART biomarkers or self-reports of currently taking ART. Finally, WLHIV were considered virally suppressed if their HIV RNA viral load was <1000 copies/mL. Women with a recent HIV infection were excluded from the ART uptake and viral suppression analyses (Supplement 1; Table S2).

### Statistical analyses

Individual-level data from each survey were pooled to calculate crude and adjusted prevalence ratios (PR) for the association between IPV and recent HIV infection, HIV testing, ART uptake and VLS. Adjusted prevalence ratios (aPR) were estimated accounting for potential confounders such as: women’s age, residence type (rural/urban), women’s marital status, women’s education, and survey-level fixed effects (survey country and year). An additional adjustment variable for the HIV recency analysis was age at sexual debut. These confounders were available from all surveys and had been previously identified as being potentially linked to both IPV and the outcomes.^44-48^ The survey-level fixed effects allowed us to control for any measured/unmeasured survey-level confounders.

Modified Poisson regression models were used to obtain the crude and adjusted PR based on Generalized Estimating Equations (GEE) with robust standard errors that accounted for the sampling design (i.e., exchangeable correlation structure with the primary sampling units as the clustering variable). Survey weights were not included in the regression^8,49^ as they are often unwarranted to obtain unbiased estimates.^50^ We used a complete case analysis since the proportion of missing observations was small for all outcomes (≲4%). Supplemental materials include information on the missing observations and the analyses of potential biases (Supplement 2). The R software (4.0.0) and the ‘geepack’ package were used for all analyses.^51^

### Sensitivity analyses

In sensitivity analyses we examined the robustness of our results by only including women testing outside of the antenatal care (ANC) in the analyses of HIV testing to examine if IPV has a differential impact by testing modality. We restricted the analysis of VLS to women on ART (i.e., conditioning on achieving this step in the cascade). To investigate if partner or couple’s characteristics confound the relationship between IPV and HIV acquisition we linked data for married or cohabiting men and women who both declared to be co-habiting. We then calculated proportions of male partners living with HIV, male partner age, partner education, partner alcohol consumption, mean partner age discrepancy and condom use at women’s most recent sex, stratified by experience of past year IPV.

All analyses were performed using secondary, de-identified data. DHS/AIS survey protocols are approved by the Internal Review Board of ICF International in Calverton (USA) and by the relevant country authorities for other surveys. Approval for data analyses was obtained from *McGill University’s Faculty of Medicine and Health Sciences Institutional Review Board* (A12-B95-21B).

## Results

### Description of included surveys

We identified 100 nationally representative surveys that included information on HIV testing (the most reported outcome), of which 64 were included in the HIV testing analysis and had data on IPV (51 DHS/AIS,5 PHIA,1 SABSSM) (Figure 1). After excluding 7 surveys, the resulting 57 surveys were conducted in 30 countries and included 280,259 unique female respondents aged 15-64 years (Supplement 3;Table S6). Women with missing data for all exposures of interest were excluded. Fifteen countries had more than one included survey and the median year of data collection was 2013. Most surveys were from Eastern Africa (51%). Only 10% of surveys had information on recent HIV infection (N_surv_=6) and 12% had data on ART uptake and VLS (N_surv_=7)(Figure 2).

**Figure 2.**
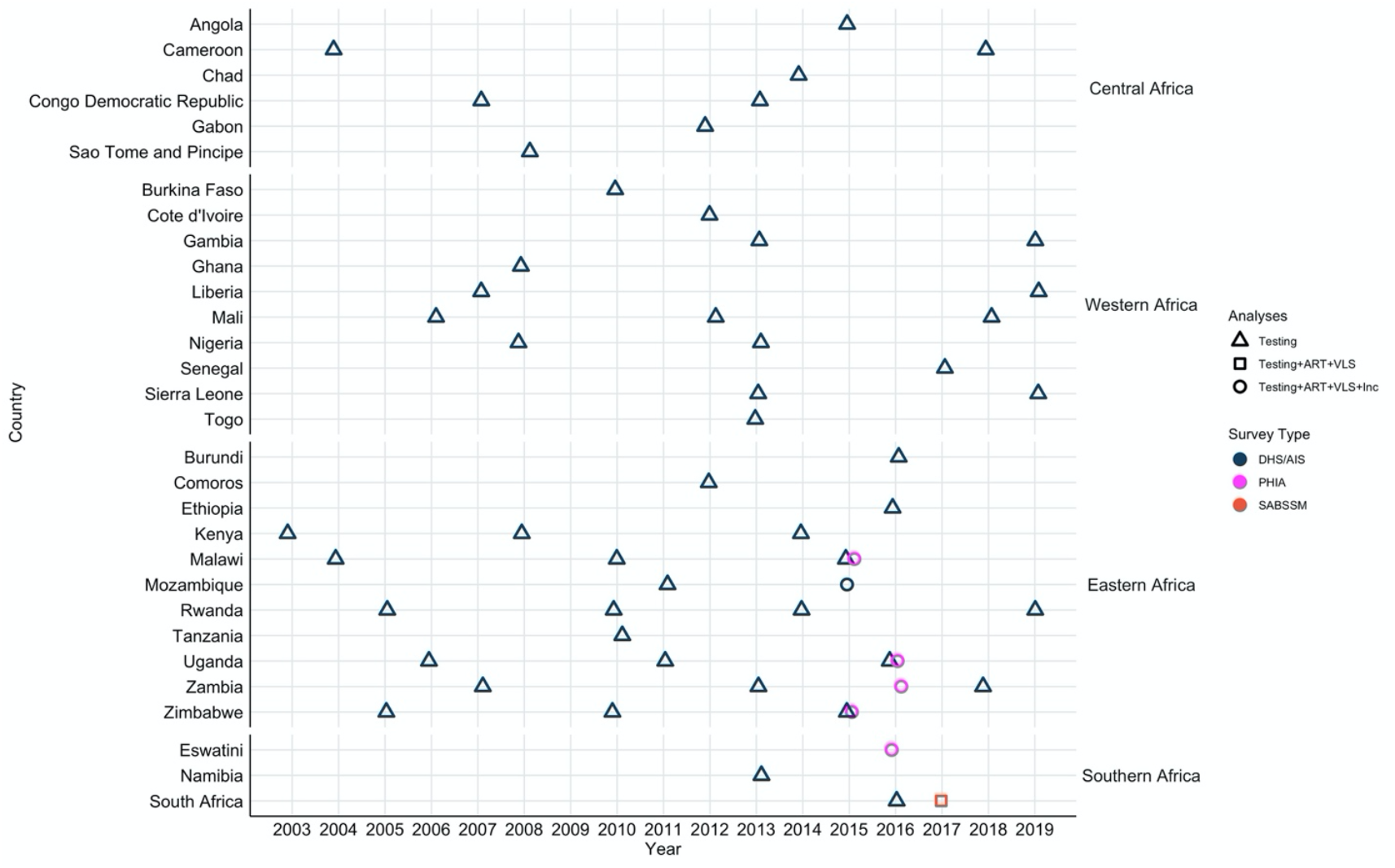
Surveys with questions on HIV and intimate partner violence (IPV), by country and year, 2000-2020. *Points represent population-based surveys conducted in sub-Saharan Africa from 2000-2020 and asking about HIV testing and intimate partner violence (IPV). Circles represent survey types. Different shapes indicate the type of analyses the surveys were included in a) only HIV testing b) HIV testing, antiretroviral (ART) uptake and viral load suppression (VLS), c) testing, ART uptake and VLS, and recent HIV infection*. (AIS=AIDS Indicator Survey; ART=antiretroviral treatment; DHS=Demographic and Health Survey; Inc=HIV incidence (recent HIV infection); PHIA=Population-based HIV Impact Assessment Survey; SABSSM=South African National HIV Prevalence, Incidence, Behaviour and Communication Survey; VLS=viral load suppression.)

### Study population

Overall, over one fifth (21%) of all ever-partnered women had experienced physical and/or sexual IPV in the past year and 29% in their lifetime. Central Africa had the highest prevalence of past year physical and/or sexual (29%) IPV, followed by Eastern Africa (23%) (Supplement 3; Table S6). Women who had experienced past year physical and/or sexual IPV were younger than those who had not, across the pooled surveys included in all the analyses (Supplement 4; Table S7-S10). Among women not living with HIV, close to half of those reporting IPV in the past year had only primary education, compared to 36% of those who did not report IPV (Supplement 4; Table S7). Most women living with HIV, irrespective of reporting IPV, had secondary education (Supplement 4;Table S8-S10).

### Recent HIV infection

Among the six surveys with information on recent infections, a total of 45 women had recently acquired HIV (Table 1). Women not living with HIV who had not experienced past year physical and/or sexual IPV had 0·5%-point higher proportion of recent HIV infections compared to those who had. The crude PR for recent HIV infections is 3·50 (95%CI: 1·63-7·51, N_surv_=6; Table 2). Adjusting for potential confounders, women who had experienced past year IPV were 3·22 times more likely (95%CI: 1·51-6·85, N_surv_=6) to have a recent HIV infection than those who had not experienced IPV. (Table 2)

**Table 1.** Unweighted proportion of women and the number of analyzed surveys with each outcome of interest: recent HIV infection, lifetime HIV testing, past year HIV testing, antiretroviral therapy (ART) uptake and viral load suppression (VLS).

| Outcome<sup>‡</sup> | Past year physical and/or sexual IPV |  |  | Lifetime physical IPV |  |  | Lifetime sexual IPV |  |  | Lifetime physical and/or sexual IPV |  |  |
| --- | --- | --- | --- | --- | --- | --- | --- | --- | --- | --- | --- | --- |
|  | Surveys (N) | Yes | No | Surveys (N) | Yes | No | Surveys (N) | Yes | No | Surveys (N) | Yes | No |
| Recent HIV infection, n (%) |  |  |  |  |  |  |  |  |  |  |  |  |
| Yes | 6 | 8<br>(0.7%) | 37<br>(0.2%) | 1 | 2<br>(0.5%) | 2<br>(0.1%) | 1 | 0<br>(0%) | 4<br>(0.2%) | 1 | 2<br>(0.5%) | 2<br>(0.1%) |
| No |  | 1,150<br>(99.3%) | 18,740<br>(99.8%) |  | 425<br>(99.5%) | 2,044<br>(99.9%) |  | 85<br>(100%) | 2,384<br>(99.8%) |  | 438<br>(99.5%) | 2,031<br>(99.9%) |
| Lifetime HIV testing, n (%) |  |  |  |  |  |  |  |  |  |  |  |  |
| Yes | 57 | 31,023<br>(52.5%) | 113,384<br>(52.4%) | 52 | 39,531<br>(53.8%) | 83,757<br>(46.2%) | 52 | 15,663<br>(55.2%) | 107,624<br>(47.6%) | 52 | 43,802<br>(54.0%) | 79,483<br>(45.8%) |
| No |  | 28,101<br>(47.5%) | 102,976<br>(47.6%) |  | 33,908<br>(46.2%) | 97,446<br>(53.8%) |  | 12,704<br>(44.8%) | 118,614<br>(52.4%) |  | 37,347<br>(46.0%) | 93,986<br>(54.2%) |
| Past year HIV testing, n (%) |  |  |  |  |  |  |  |  |  |  |  |  |
| Yes | 57 | 16,392<br>(27.8%) | 57,996<br>(26.9%) | 52 | 20,292<br>(27.7%) | 43,451<br>(24.0%) | 52 | 8,240<br>(29.1) | 55,506<br>(24.6) | 52 | 22,555<br>(27.9%) | 41,189<br>(23.8%) |
| No |  | 42,601<br>(72.2%) | 157,969<br>(73.1%) |  | 52,993<br>(72.3%) | 137,456<br>(76.0%) |  | 20,080<br>(70.9%) | 170,329<br>(75.4%) |  | 58,429<br>(72.1%) | 131,995<br>(76.2%) |
| ART uptake, n (%) |  |  |  |  |  |  |  |  |  |  |  |  |
| Yes | 7 | 416<br>(64.2%) | 3,717<br>(71.3%) | 2 | 388<br>(64.8%) | 675<br>(60.5%) | 2 | 68<br>(66.7%) | 996<br>(61.7%) | 2 | 402<br>(64.9%) | 661<br>(60.3%) |
| No |  | 232<br>(35.8%) | 1,498<br>(28.7%) |  | 211<br>(35.2%) | 441<br>(39.5%) |  | 34<br>(33.3%) | 618<br>(38.3%) |  | 217<br>(35.1%) | 435<br>(39.7%) |
| VLS, n (%) |  |  |  |  |  |  |  |  |  |  |  |  |
| Yes | 7 | 375<br>(56.7%) | 3,506<br>(67.3%) | 2 | 348<br>(57.0%) | 645<br>(57.3%) | 2 | 60<br>(60.6%) | 934<br>(57.0%) | 2 | 362<br>(57.4%) | 631<br>(57.1%) |
| No |  | 286<br>(43.3%) | 1,700<br>(32.7%) |  | 262<br>(43.0%) | 481<br>(42.7%) |  | 39<br>(39.4%) | 704<br>(43.0%) |  | 269<br>(42.6%) | 474<br>(42.9%) |

ART=antiretroviral therapy; IPV=intimate partner violence; Surv.=survey; VLS=viral load suppression.
‡Denominators for these proportions are the total number of women with or without each of the exposures of interest: past year physical and/or sexual IPV, lifetime physical IPV, lifetime sexual IPV, lifetime physical and/or sexual IPV.

**Table 2.**
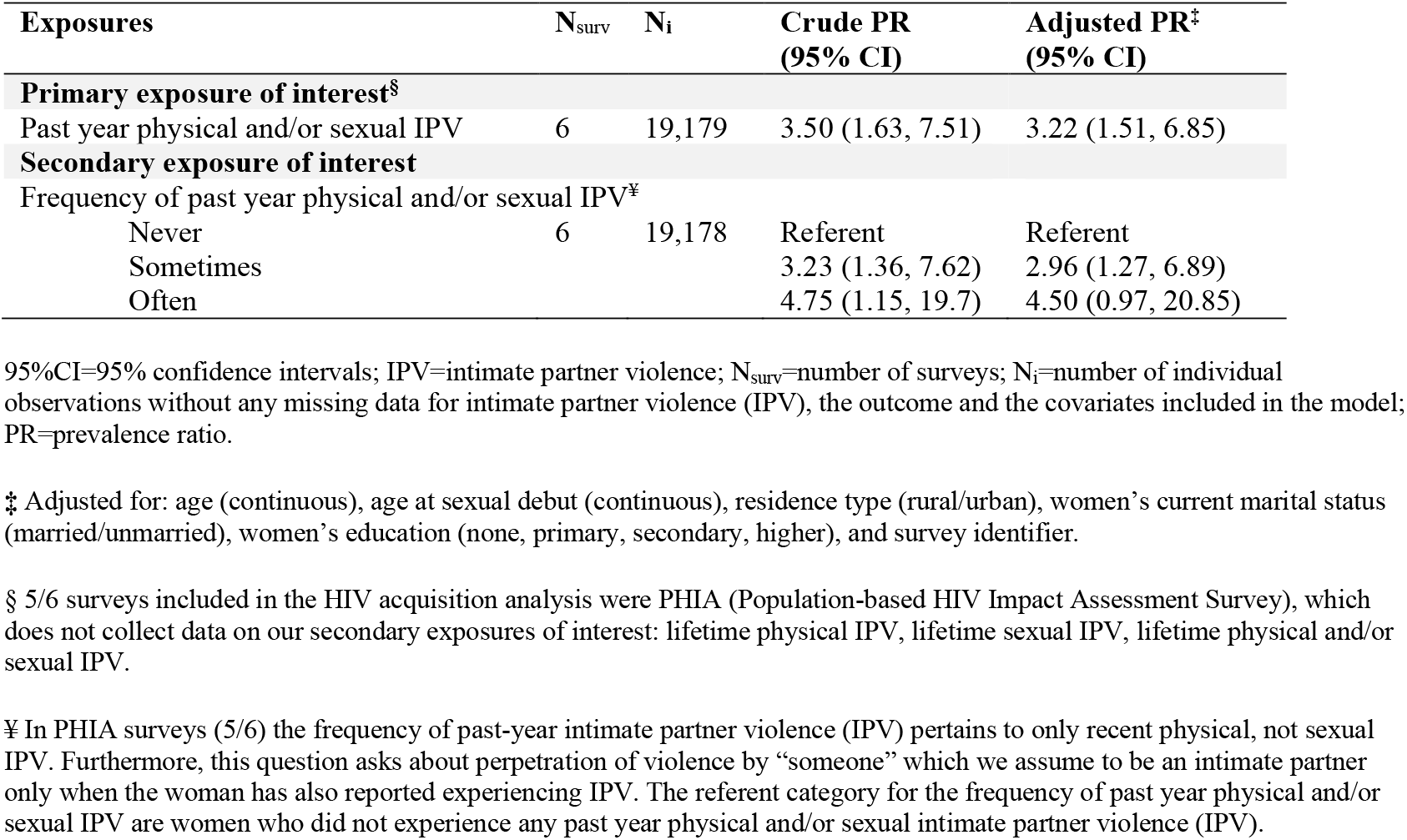
Crude and adjusted prevalence ratios of HIV recent infection among women experiencing past-year physical and/or sexual intimate partner violence (IPV) compared to women not experiencing this type of IPV.

As a robustness check, we examined the HIV status of the cohabiting partners, partners’ age discrepancy, partner education, partner alcohol consumption, and condom use at women’s last sex as potential confounders in the six surveys included in the recent HIV infection analysis (Supplement 5; Table S11-S14). Partner age discrepancy and partner alcohol consumption varied between women having experienced past year IPV and those who had not; however the effect estimates are robust to confounding by these variables.

### HIV testing

Self-reports of HIV testing in the past year were similar between women who had experienced IPV and those who had not. About a quarter of women in both groups had been tested in the past year (28% and 27% among those who experienced IPV and those who had not, respectively) and nearly half of all women (regardless of experience of IPV in the past year) ever tested for HIV (Table 1).

The crude PR of past year physical and/or sexual IPV on recent HIV testing was 0·98 (95%CI: 0·97-0·99). Adjusting for potential confounders, experience of past year IPV had no effect on recent HIV testing (aPR=0·99, 95%CI: 0·98-1·01, N_surv_=57). Women experiencing lifetime physical and/or sexual IPV were 2% more likely to report lifetime testing (aPR= 1·02, 95%CI: 1·02-1·03, N_surv_=52) (Table 3).

**Table 3.**
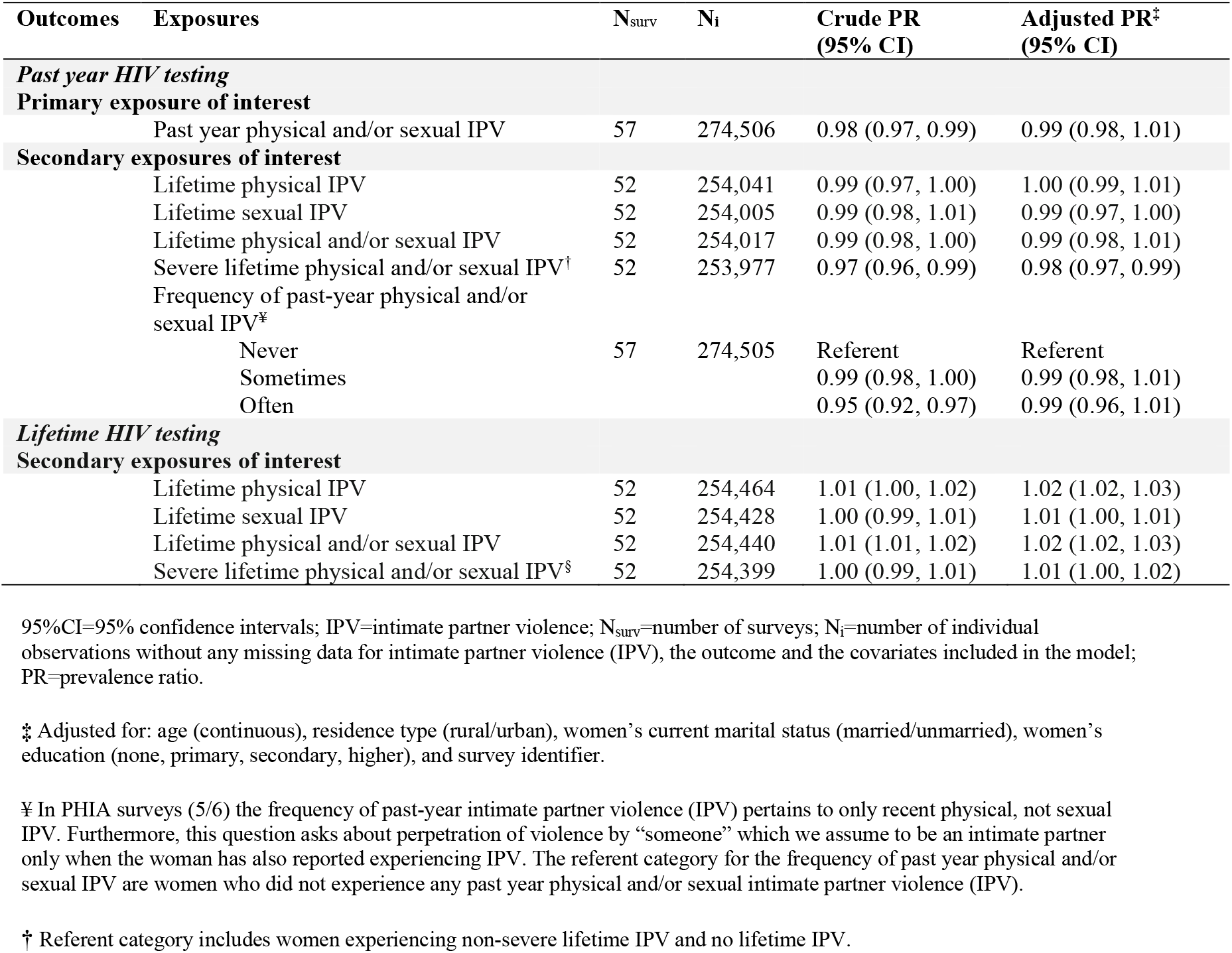
Crude and adjusted prevalence ratios of lifetime and past year HIV testing among women experiencing different intimate partner violence (IPV) categories compared to women not experiencing a specific type of IPV.

Similar results were obtained in sensitivity analyses where we excluded women who reported testing at ANC. Experience of lifetime physical and/or sexual IPV was associated with a small increase in lifetime HIV testing among women who tested outside of the ANC (aPR=1·04, 95%CI: 1·03-1·05, N_surv_=52; Supplement 6;Table S15).

### Antiretroviral therapy uptake

WLHIV who had reported past year physical and/or sexual IPV compared to those who had not had 7·1%-point lower ART uptake (Table 1). The crude PR for ART uptake was 0·90 (95%CI: 0·85-0·96, N_surv_=7). After adjustments, women who had reported past year IPV were 4% less likely to be on ART, compared to those who had not (aPR=0·96, 95%CI: 0·90-1·03, N_surv_=7), but we cannot rule out the absence of an effect. Severe lifetime IPV slightly increased the strength of the association with poor ART uptake (aPR=0·94, 95%CI: 0·84-1·05, N_surv_=2), though this analysis was restricted to two surveys with information which decreased the precision of the estimate (Table 4).

**Table 4.** Crude and adjusted prevalence ratios of antiretroviral (ART) uptake among women living with HIV (WLHIV) experiencing different intimate partner violence (IPV) categories compared to WLHIV not experiencing a specific type of IPV

| Exposures | N<sub>surv</sub> | N<sub>i</sub><sup>§</sup> | Crude PR<br>(95% CI) | Adjusted PR<sup>‡</sup><br>(95% CI) |
| --- | --- | --- | --- | --- |
| **Primary exposure of interest** |  |  |  |  |
| Past year physical and/or sexual IPV | 7 | 5,629 | 0.90 (0.85, 0.96) | 0.96 (0.9, 1.03) |
| **Secondary exposures of interest** |  |  |  |  |
| Lifetime physical IPV | 2 | 1,569 | 1.06 (0.98, 1.14) | 1.00 (0.93, 1.08) |
| Lifetime sexual IPV | 2 | 1,570 | 1.08 (0.94, 1.24) | 1.05 (0.91, 1.21) |
| Lifetime physical and/or sexual IPV | 2 | 1,569 | 1.06 (0.98, 1.14) | 1.01 (0.94, 1.09) |
| Severe lifetime physical and/or sexual IPV<sup>†</sup> | 2 | 1,568 | 0.93 (0.83, 1.04) | 0.94 (0.84, 1.05) |
| Frequency of past-year physical and/or sexual IPV<sup>¥</sup> |  |  |  |  |
| Never | 7 | 5,629 | Referent | Referent |
| Sometimes |  |  | 0.91 (0.86, 0.97) | 0.96 (0.90, 1.03) |
| Often |  |  | 0.84 (0.71, 0.99) | 0.97 (0.82, 1.16) |

95%CI=95% confidence intervals; ART=antiretroviral treatment; IPV=intimate partner violence; N<sub>surv</sub>=number of surveys; N<sub>i</sub>=number of individual observations without any missing data for intimate partner violence (IPV), the outcome and the covariates included in the model; PR=prevalence ratio; WLHIV=women living with HIV.
<sup>‡</sup> Adjusted for: age (continuous), residence type (rural/urban), women's current marital status (married/unmarried), women's education (none, primary, secondary, higher), and survey identifier.
<sup>¥</sup> In PHIA surveys (5/6) the frequency of past-year intimate partner violence (IPV) pertains to only recent physical, not sexual IPV. Furthermore, this question asks about perpetration of violence by "someone" which we assume to be an intimate partner only when the woman has also reported experiencing IPV. The referent category for the frequency of past year physical and/or sexual IPV are women who did not experience any past year physical and/or sexual intimate partner violence (IPV).
<sup>§</sup> 0.7% of women were removed because they had a recent HIV infection and did not have time to be diagnosed and linked to treatment.
<sup>†</sup> Referent category includes women experiencing non-severe lifetime IPV and no lifetime IPV.

### Viral load suppression

WLHIV who had experienced past year physical and/or sexual IPV compared to those who had not had 10·6%-point lower VLS (Table 1). The crude PR for VLS was 0·85 (95%CI:0·79-0·91, N_surv_=7). After adjusting for confounders, women who had experienced past year IPV were 9% less likely to be virally suppressed, compared to those who had not (aPR=0·91, 95%CI: 0·85-0·98, N_surv_=7; Table 5). Lifetime physical and/or sexual IPV had an adverse effect on VLS as well, though the confidence interval includes the null (aPR=0·94, 95%CI:0·86-1·02, N_surv_=2).

**Table 5.** Crude and adjusted prevalence ratios of viral load suppression (VLS) among women living with HIV (WLHIV) experiencing different intimate partner violence (IPV) categories compared to WLHIV not experiencing a specific type of IPV.

| Exposures | N<sub>surv</sub> | N<sub>i</sub><sup>§</sup> | Crude PR<br>(95% CI) | Adjusted PR<sup>‡</sup><br>(95% CI) |
| --- | --- | --- | --- | --- |
| **Primary exposure of interest** |  |  |  |  |
| Past year physical and/or sexual IPV | 7 | 5,627 | 0.85 (0.79, 0.91) | 0.91 (0.85, 0.98) |
| **Secondary exposures of interest** |  |  |  |  |
| Lifetime physical IPV | 2 | 1,583 | 0.99 (0.91, 1.07) | 0.93 (0.85, 1.01) |
| Lifetime sexual IPV | 2 | 1,584 | 1.08 (0.91, 1.27) | 1.04 (0.88, 1.23) |
| Lifetime physical and/or sexual IPV | 2 | 1,583 | 1.00 (0.92, 1.08) | 0.94 (0.86, 1.02) |
| Severe lifetime physical and/or sexual IPV<sup>†</sup> | 2 | 1,582 | 0.99 (0.88, 1.12) | 1.01 (0.90, 1.14) |
| Frequency of past-year physical and/or sexual IPV<sup>¶</sup> |  |  |  |  |
| Never | 7 | 5,627 | Referent | Referent |
| Sometimes |  |  | 0.86 (0.8, 0.93) | 0.91 (0.84, 0.98) |
| Often |  |  | 0.74 (0.61, 0.91) | 0.89 (0.73, 1.09) |

95%CI=95% confidence intervals; IPV=intimate partner violence; PR=prevalence ratio; N<sub>surv</sub>=number of surveys; N<sub>i</sub>=number of individual observations without any missing data for intimate partner violence (IPV), the outcome and the covariates included in the model; VLS=viral load suppression; WLHIV=women living with HIV.
<sup>‡</sup> Adjusted for: age (continuous), residence type (rural/urban), women's current marital status (married/unmarried), women's education (none, primary, secondary, higher), and survey identifier.
<sup>¶</sup> In PHIA surveys (5/6) the frequency of past-year intimate partner violence (IPV) pertains to only recent physical, not sexual IPV. Furthermore, this question asks about perpetration of violence by "someone" which we assume to be an intimate partner only when the woman has also reported experiencing IPV. The referent category for the frequency of past year physical and/or sexual IPV are women who did not experience any past year physical and/or sexual intimate partner violence (IPV).
<sup>§</sup> 0.7% of women were removed because they had a recent HIV infection and did not have time to be diagnosed and linked to treatment.
<sup>†</sup> Referent category includes women experiencing non-severe lifetime IPV and no lifetime IPV.

We further looked at the effect of IPV on VLS, restricting to women on ART. The effect size estimate between past year IPV and VLS among WLHIV on ART was smaller. WLHIV on ART who had experienced past year IPV were 6% less likely to have VLS compared to women who had not (aPR=0·94, 95%CI: 0·90-0·99, N_surv_=7; Supplement 6; Table S16). Higher frequency of IPV in the past year was associated with a 14% reduction in the likelihood of VLS although the estimates were imprecise (aPR=0·86, 95%CI: 0·72-1·02, N_surv_=7).

## Discussion

IPV is a human right violation, a global health concern, and our study shows that it could contribute to HIV epidemics. In our pooled analysis of population-based surveys, women who had experienced physical and/or sexual IPV over the past year were over 3 times more likely to have acquired a recent HIV infection compared to those who had not. Although we did not find evidence of the impact of past year IPV on HIV testing, WLHIV who experienced physical and/or sexual IPV in the past year were 9% less likely to be virally suppressed. In line with the *United Nations 2021 Political Declaration* to end gender inequalities perpetuating the HIV/AIDS epidemic, the available evidence^9,10^ suggest that elimination of IPV could be partly responsible for reductions in new HIV infections and improvements viral load suppresion.^52^

In sub-Saharan Africa, the links between IPV and HIV have previously been studied using HIV prevalence as the outcome.^5-8,49^ Such study designs make it difficult to disentangle temporality between experience of IPV and HIV acquisition. We leveraged recent infection assays that were recently deployed in large population-based surveys such as PHIA, as a proxy for HIV incidence. Our results align with previous cohort studies, but our effect size estimates are larger.^6,9,10^ Longitudinal studies in South Africa and Uganda show that women who had been subjected to physical or sexual IPV had 1.5 times the HIV incidence compared to those who had not.^9,10^ Larger effect size in our study may be attributable to the exposure-driven selection bias in cohort studies and/or generalizability of effect size estimates from randomized controlled trials and cohort studies.^9,10^

Pathways through which IPV can affect HIV acquisition are complex and multifaceted. While the most direct path is through sexual assault, a growing body of evidence suggests that at the population level, structural factors surrounding IPV play a larger role.^53,54^ Men who perpetrate IPV may be more likely to have a high number of concurrent sexual partners, use condoms inconsistently, and use substances, and thus are more likely to be living with HIV which in turn could lead to HIV transmission to their partners.^54-58^ Crude, descriptive evidence from our study suggests that the proportion of partners who are living with HIV and condom use at last sex are comparable between women experiencing and not experiencing IPV in the past year. Partner age discrepancy and partner alcohol consumption varied between women who had experienced IPV and those who had not, but the effect estimates remained robust to confounding by these covariates.

Knowledge of HIV status among WLHIV –the first step in the cascade– can be a key bottleneck in the HIV care cascade. In our study, IPV did not impact HIV testing, even after excluding women who had tested at ANC (as HIV testing at ANC achieved high coverage). Overall, evidence regarding the effect of IPV on HIV testing is mixed. Some studies suggested lower rates of HIV testing due to a fear of violent reaction from one’s partner if the HIV test comes back positive.^20,25,59-61^ Most studies from low-and middle-income countries found no relationship, or even a positive relationship between IPV and HIV testing, which might be due to a higher self-perceived risk among the IPV survivor.^15,16,18,62,63^ This is corroborated in our study, where lifetime experience of physical and/or sexual IPV is associated with a small increase in lifetime HIV testing, which might speak to the cumulative nature of lifetime experience of IPV and associated high self-perceived risk. Null results could also be reflective of the unprecedented scale-up of HIV-testing in sub-Saharan Africa in the past decade.^37^

Our results suggest that women who had experienced past year IPV were less likely to be on ART, compared to those who had not, though our results were not conclusive. Most existing studies conducted in low- and middle-income countries look at the effects of IPV on ART adherence. Those that assessed current ART use did not uncover relationships between the two, which aligns with our results.^13^ However, we found that IPV was adversely associated with VLS, even among women who are on ART, which implies that ART adherence could be the bottleneck in WLHIV’s success in the HIV care cascade. Pathways through which IPV affects ART uptake and adherence are complex. Some women might hide their HIV status due to fear of their partner’s reaction, making it difficult to enroll in HIV care and adhere to treatment.^14,64-66^ IPV could restrict women’s access to healthcare through male control of resources, and freedoms of mobility and decision-making.^67-69^

Our results should be interpreted considering the study’s limitations. First, all surveys depend on self-reports of IPV and might be subject to under-reporting due to their sensitive nature.^70^ However, the surveys used appropriate measures to ensure confidentiality. HIV testing was also self-reported, though evidence shows that self-reported HIV-testing histories are generally robust.^71^ Second, some of the included surveys slightly differed in terms of their question wordings on IPV (e.g., frequency of past-year violence was a continuous variable and had to be categorized). Nevertheless, questions were all acts-based and modified from the *Conflict Tactics Scales*.^38^ Third, we did not include emotional violence in the definition of intimate partner violence, due to a lack of consensus on how to define, conceptualize, and measure this construct cross-culturally.^72^ Fourth, few surveys had information on recent HIV infection, ART uptake and VLS as compared to HIV testing histories. However, pooling all data sources allowed us to improve the precision of our estimates. Fifth, we used cross-sectional survey data and reverse causality remains a possibility. This limitation was alleviated, however, by restricting our main exposure to recent IPV, examining recent HIV infection, and ART uptake and VLS at the time of the interview. Sixth, non-response could threaten the validity of our results, leading to selection bias and potential underestimation of the effect size measure (Supplement 2). Finally, our descriptive analyses of male partner characteristics were based on a small sample, and we cannot rule out residual confounding.

Strengths of our study include a comprehensive analysis of available population-based surveys with information on IPV and HIV. Second, our large sample size allowed us to estimate the adjusted prevalence ratios for the relationships between IPV, recent HIV infection and each stage of the HIV treatment and care cascade. Finally, we conducted several sensitivity analyses to assess the robustness of our findings.

Our results have important policy implications for HIV prevention and care delivery in high HIV burden settings. At a service delivery-level, healthcare provider training should include IPV-sensitive topics such as probing women in a nonjudgmental way to safely disclose their experience of IPV. This could identify women at higher risk of disengagement from care who can subsequently be linked to women-centric HIV services. Emergence of novel, patient-focused HIV service delivery platforms, also known as “*differentiated service delivery*” models could incorporate women-only community adherence groups or safe community-based medication pick-up points.^73^

At a macro level, harmful gender norms remain worldwide such that a large proportion of youth still agree that a husband is sometimes justified in beating his wife.^32^ Systemic work is required to eradicate such attitudes and to mitigate their impact on HIV outcomes among women, such as HIV educational campaigns for youth that have a strong gender equity component.^74^ National legislative systems must provide frameworks for these structural changes. Although in 103 countries, criminal penalties exist for IPV, their implementation varies.^32^ Such laws must be better enacted and enforced as the national legal environments often deepen gender inequality and undermine national HIV responses.

In conclusion, IPV could have important adverse effects on HIV epidemics by contributing to HIV acquisition risks and decreasing VLS among WLHIV. Efforts to prevent intimate partner violence and violence against women more broadly should be included in HIV control programs and indicators of IPV collected and monitored as part of HIV surveillance. The IPV/HIV syndemic needs recognition by governments and communities if we are to eliminate IPV and reduce HIV risk among women.

## Data Availability

All data produced in the present study are available upon reasonable request to the authors

## Contributions

SK and MMG conceived of the study. SK performed the analyses. MMG and DP contributed to the study methods and reviewing and editing the manuscript. SK wrote the initial draft of the manuscript. DP, MCB, SD, JWE, FM, LS, LSC, HS, RW and MMG helped interpret the results and provided valuable feedback on the policy implications of the study. All authors contributed to and approved the final manuscript.

## Acknowledgments

We are grateful and indebted to the survey teams who made their data publicly available and to all survey participants.

## Data sharing

Data used in the study are publicly available for investigators who submit an abstract and a data analysis plan as part of *Demographic and Health Surveys* (DHS), *AIDS Indicator Surveys* (AIS) (https://dhsprogram.com/Data/), *Population-based HIV Impact Assessment* (PHIA) (https://phia-data.icap.columbia.edu/datasets) and *South Africa National HIV Prevalence, Incidence, Behavior and Communication Survey* (SABSSM) (http://datacuration.hsrc.ac.za/search/browse/alpha/S).

Analysis code and clean data that support the findings of the study are available upon request.

## References

1. Garcia-Moreno C. Violence Against Women: International Perspectives. American Journal of Preventive Medicine 2000; 19(4): 330–3.

2. Garcia-Moreno C, Heise L, Jansen HA, Ellsberg M, Watts C. Public health. Violence against women. Science 2005; 310(5752): 1282–3.

3. Garcia-Moreno C, Watts C. Violence against women: its importance for HIV/AIDS. AIDS 2000; 14 Suppl 3: S253–65.

4. Martin SL, Curtis S. Gender-based violence and HIV/AIDS: recognising links and acting on evidence. Lancet 2004; 363(9419): 1410–1.

5. Kouyoumdjian FG, Findlay N, Schwandt M, Calzavara LM. A systematic review of the relationships between intimate partner violence and HIV/AIDS. PLoS One 2013; 8(11): e81044.

6. Li Y, Marshall CM, Rees HC, Nunez A, Ezeanolue EE, Ehiri JE. Intimate partner violence and HIV infection among women: a systematic review and meta-analysis. J Int AIDS Soc 2014; 17: 18845.

7. Campbell JC, Baty ML, Ghandour RM, Stockman JK, Francisco L, Wagman J. The intersection of intimate partner violence against women and HIV/AIDS: a review. Int J Inj Contr Saf Promot 2008; 15(4): 221–31.

8. Durevall D, Lindskog A. Intimate partner violence and HIV in ten sub-Saharan African countries: what do the Demographic and Health Surveys tell us? Lancet Glob Health 2015; 3(1): e34–43.

9. Kouyoumdjian FG, Calzavara LM, Bondy SJ, et al. Risk factors for intimate partner violence in women in the Rakai Community Cohort Study, Uganda, from 2000 to 2009. BMC Public Health 2013; 13(566).

10. Jewkes RK, Dunkle K, Nduna M, Shai N. Intimate partner violence, relationship power inequity, and incidence of HIV infection in young women in South Africa: a cohort study. Lancet 2010; 376(9734): 41–8.

11. Zablotska IB, Gray RH, Koenig MA, et al. Alcohol use, intimate partner violence, sexual coercion and HIV among women aged 15-24 in Rakai, Uganda. AIDS Behav 2009; 13(2): 225–33.

12. Were E, Curran K, Delany-Moretlwe S, et al. A prospective study of frequency and correlates of intimate partner violence among African heterosexual HIV serodiscordant couples. AIDS 2011; 25(16): 2009–18.

13. Hatcher AM, Smout EM, Turan JM, Christofides N, Stockl H. Intimate partner violence and engagement in HIV care and treatment among women: a systematic review and meta-analysis. AIDS 2015; 29(16): 2183–94.

14. Leddy AM, Weiss E, Yam E, Pulerwitz J. Gender-based violence and engagement in biomedical HIV prevention, care and treatment: a scoping review. BMC Public Health 2019; 19(1): 897.

15. Nelson KA, Ferrance JL, Masho SW. Intimate partner violence, consenting to HIV testing and HIV status among Zambian women. Int J STD AIDS 2016; 27(10): 832–9.

16. Nikolova SP, Small E, Mengo C. Components of resilience in gender: a comparative analysis of HIV outcomes in Kenya. Int J STD AIDS 2015; 26(7): 483–95.

17. Lyons CE, Grosso A, Drame FM, et al. Physical and Sexual Violence Affecting Female Sex Workers in Abidjan, Cote d’Ivoire: Prevalence, and the Relationship with the Work Environment, HIV, and Access to Health Services. J Acquir Immune Defic Syndr 2017; 75(1): 9–17.

18. Conroy A, Leddy A, Johnson M, Ngubane T, van Rooyen H, Darbes L. ‘I told her this is your life’: relationship dynamics, partner support and adherence to antiretroviral therapy among South African couples. Cult Health Sex 2017; 19(11): 1239–53.

19. Kiarie JN, Farquhar C, Richardson BA, et al. Domestic violence and prevention of mother-to-child transmission of HIV-1. AIDS 2006; 20(13): 1763–9.

20. Mohammed BH, Johnston JM, Harwell JI, Yi H, Tsang KW, Haidar JA. Intimate partner violence and utilization of maternal health care services in Addis Ababa, Ethiopia. BMC Health Serv Res 2017; 17(1): 178.

21. Turan JM, Bukusi EA, Onono M, Holzemer WL, Miller S, Cohen CR. HIV/AIDS stigma and refusal of HIV testing among pregnant women in rural Kenya: results from the MAMAS Study. AIDS Behav 2011; 15(6): 1111–20.

22. Kidman R, Violari A. Dating Violence Against HIV-Infected Youth in South Africa: Associations With Sexual Risk Behavior, Medication Adherence, and Mental Health. J Acquir Immune Defic Syndr 2018; 77(1): 64–71.

23. Hampanda K. Intimate Partner Violence Against HIV-Positive Women is Associated with Sub-Optimal Infant Feeding Practices in Lusaka, Zambia. Matern Child Health J 2016; 20(12): 2599–606.

24. Biomndo BC, Bergmann A, Lahmann N, Atwoli L. Intimate partner violence is a barrier to antiretroviral therapy adherence among HIV-positive women: Evidence from government facilities in Kenya. PLoS One 2021; 16(4): e0249813.

25. Hatcher AM, Stöckl H, Christofides N, et al. Mechanisms linking intimate partner violence and prevention of mother-to-child transmission of HIV: A qualitative study in South Africa. Soc Sci Med 2016; 168: 130–9.

26. Merrill KG, Campbell JC, Decker MR, et al. Past-Year Violence Victimization is Associated with Viral Load Failure Among HIV-Positive Adolescents and Young Adults. AIDS Behav 2021; 25(5): 1373–83.

27. Hatcher AM, Brittain K, Phillips TK, Zerbe A, Abrams EJ, Myer L. Longitudinal association between intimate partner violence and viral suppression during pregnancy and postpartum in South African women. AIDS 2021; 35(5): 791–9.

28. UNAIDS. UNAIDS Data 2021, 2021.

29. UNAIDS. Global HIV Statistics: Fact Sheet, 2021.

30. UNAIDS. Fast-Track: Ending the AIDS Epidemic by 2030, 2014.

31. Sardinha L M-GM, Stockl H, Meyer S, Garcia-Moreno C. Global, regional and national prevalence estimates of physical and/or sexual intimate partner violence against women, 2018. Lancet 2022; 399(10327): P803–13.

32. UNAIDS. Global AIDS update: Confronting inequalities, 2021.

33. Schaefer R, Gregson S, Fearon E, Hensen B, Hallett TB, Hargreaves JR. HIV prevention cascades: A unifying framework to replicate the successes of treatment cascades. Lancet HIV 2019; 6(1): e60–e6.

34. Hampanda KM. Intimate partner violence and HIV-positive women’s non-adherence to antiretroviral medication for the purpose of prevention of mother-to-child transmission in Lusaka, Zambia. Soc Sci Med 2016; 153: 123–30.

35. Wechsberg WM, van der Horst C, Ndirangu J, et al. Seek, test, treat: substance-using women in the HIV treatment cascade in South Africa. Addict Sci Clin Pract 2017; 12(1): 12.

36. Stockl H, Sardinha L, Maheu-Giroux M, Meyer SR, Garcia-Moreno C. Physical, sexual and psychological intimate partner violence and non-partner sexual violence against women and girls: a systematic review protocol for producing global, regional and country estimates. BMJ Open 2021; 11(8): e045574.

37. Giguere K, Eaton JW, Marsh K, et al. Trends in knowledge of HIV status and efficiency of HIV testing services in sub-Saharan Africa, 2000-20: a modelling study using survey and HIV testing programme data. Lancet HIV 2021; 8(5): e284–e93.

38. Straus M. The Revised Conflict Tactics Scale. Journal of Family Issues 1996; 17(3): 286–316.

39. Kishor S. Domestic violence measurement in the demographic and health surveys: The history and the challenges. Geneva, Switzerland, 2005.

40. Maheu-Giroux M SL, Stöckl H, Meyer S, Godin A, Alexander M, García-Moreno C. [Preprint] A framework to model global, regional, and national estimates of intimate partner violence. 2022.

41. Laeyendecker O, Gray RH, Grabowski MK, et al. Validation of the Limiting Antigen Avidity Assay to Estimate Level and Trends in HIV Incidence in an A/D Epidemic in Rakai, Uganda. AIDS Res Hum Retroviruses 2019; 35(4): 364–7.

42. WHO. HIV Incidence Meaasurement and Data Use. Boston, 2018.

43. Mitchell KM, Maheu-Giroux M, Dimitrov D, et al. How can progress towards Ending the HIV Epidemic in the United States be monitored? Clin Infect Dis 2021.

44. Hallett TB, Lewis JJ, Lopman BA, et al. Age at first sex and HIV infection in rural Zimbabwe. Stud Fam Plann 2007; 38(1): 1–10.

45. Kouyoumdjian F. Intimate Partner Violence as a Risk Factor for Incident HIV Infection in Women in Rakai, Uganda. Toronto: University of Toronto; 2014.

46. Shamu S, Abrahams N, Temmerman M, Musekiwa A, Zarowsky C. A systematic review of African studies on intimate partner violence against pregnant women: prevalence and risk factors. PLoS One 2011; 6(3): e17591.

47. Auvert B, Buve A, Ferry B, et al. Ecological and individual level analysis of risk factors for HIV infection in four urban populations in sub-Saharan Africa with different levels of HIV infection. AIDS 2001; 15: S15–30.

48. Nabaggala MS, Reddy T, Manda S. Effects of rural-urban residence and education on intimate partner violence among women in Sub-Saharan Africa: a meta-analysis of health survey data. BMC Womens Health 2021; 21(1): 149.

49. Harling G, Msisha W, Subramanian SV. No association between HIV and intimate partner violence among women in 10 developing countries. PLoS One 2010; 5(12): e14257.

50. Solon GH, S and Wooldridge J. [Working paper] What Are We Weighting For? In: Research NBoE, editor.; 2013.

51. Højsgaard S. Package ‘geepack’, 2022.

52. Wagman JA, Gray RH, Campbell JC, et al. Effectiveness of an integrated intimate partner violence and HIV prevention intervention in Rakai, Uganda: analysis of an intervention in an existing cluster randomised cohort. Lancet Glob Health 2015; 3(1): e23–33.

53. McLean I, Roberts SA, White C, Paul S. Female genital injuries resulting from consensual and non-consensual vaginal intercourse. Forensic Sci Int 2011; 204(1-3): 27–33.

54. Dunkle KL, Decker MR. Gender-based violence and HIV: reviewing the evidence for links and causal pathways in the general population and high-risk groups. Am J Reprod Immunol 2013; 69: 20–6.

55. Decker MR, Seage GR, 3rd, Hemenway D, et al. Intimate partner violence functions as both a risk marker and risk factor for women’s HIV infection: findings from Indian husband-wife dyads. J Acquir Immune Defic Syndr 2009; 51(5): 593–600.

56. Jewkes R, Sikweyiya Y, Morrell R, Dunkle K. The relationship between intimate partner violence, rape and HIV amongst South African men: a cross-sectional study. PLoS One 2011; 6(9): e24256.

57. Jewkes R, Dunkle K, Nduna M, et al. Factors associated with HIV sero-status in young rural South African women: connections between intimate partner violence and HIV. Int J Epidemiol 2006; 35(6): 1461–8.

58. Silverman JG, McCauley HL, Decker MR, Miller E, Reed E, Raj A. Coercive forms of sexual risk and associated violence perpetrated by male partners of female adolescents. Perspect Sex Reprod Health 2011; 43(1): 60–5.

59. Pearlman DN, Averbach AR, Zierler S, Cranston K. Disparities in prenatal HIV testing: evidence for improving implementation of CDC screening guidelines. J Natl Med Assoc 2005; 97(7): 44S–51S.

60. Etudo O, Metheny N, Stephenson R, Kalokhe AS. Intimate partner violence is linked to less HIV testing uptake among high-risk, HIV-negative women in Atlanta. AIDS Care 2017; 29(8): 953–6.

61. Nasrullah M, Oraka E, Breiding MJ, Chavez PR. HIV testing and intimate partner violence among non-pregnant women in 15 US states/territories: findings from behavioral risk factor surveillance system survey data. AIDS Behav 2013; 17(7): 2521–7.

62. Satyanarayana VA, Chandra PS, Vaddiparti K, Benegal V, Cottler LB. Factors influencing consent to HIV testing among wives of heavy drinkers in an urban slum in India. AIDS Care 2009; 21(5): 615–21.

63. Raj A, Silverman JG, Amaro H. Abused women report greater male partner risk and gender-based risk for HIV: findings from a community-based study with Hispanic women. AIDS Care 2004; 16(4): 519–29.

64. Sullivan KA, Messer LC, Quinlivan EB. Substance abuse, violence, and HIV/AIDS (SAVA) syndemic effects on viral suppression among HIV positive women of color. AIDS Patient Care STDS 2015; 29: S42–8.

65. Espino SR, Fletcher J, Gonzalez M, Precht A, Xavier J, Matoff-Stepp S. Violence screening and viral load suppression among HIV-positive women of color. AIDS Patient Care STDS 2015; 29(1): S36–41.

66. Machtinger EL, Haberer JE, Wilson TC, Weiss DS. Recent trauma is associated with antiretroviral failure and HIV transmission risk behavior among HIV-positive women and female-identified transgenders. AIDS Behav 2012; 16(8): 2160–70.

67. Maman S, Mbwambo JK, Hogan NM, et al. HIV-positive women report more lifetime partner violence: findings from a voluntary counseling and testing clinic in Dar es Salaam, Tanzania. Am J Public Health 2002; 92(8): 1331–7.

68. Khan MN, Islam MM. Women’s attitude towards wife-beating and its relationship with reproductive healthcare seeking behavior: A countrywide population survey in Bangladesh. PLoS One 2018; 13(6): e0198833.

69. Daftary A, Padayatchi N, Padilla M. HIV testing and disclosure: a qualitative analysis of TB patients in South Africa. AIDS Care 2007; 19(4): 572–7.

70. Ellsberg M, Heise L, Peña R, Agurto S, Winkvist A. Researching domestic violence against women: methodological and ethical considerations. Stud Fam Plann 2001; 32(1): 1–16.

71. Xia Y, Milwid RM, Godin A, et al. Accuracy of self-reported HIV-testing history and awareness of HIV-positive status in four sub-Saharan African countries. AIDS 2021; 35(3): 503–10.

72. Heise L, Pallitto C, Garcia-Moreno C, Clark CJ. Measuring psychological abuse by intimate partners: Constructing a cross-cultural indicator for the Sustainable Development Goals. SSM Popul Health 2019; 9: 100377.

73. Roy M, Bolton Moore C, Sikazwe I, Holmes CB. A Review of Differentiated Service Delivery for HIV Treatment: Effectiveness, Mechanisms, Targeting, and Scale. Curr HIV/AIDS Rep 2019; 16(4): 324–34.

74. Sharma V, Leight J, Verani F, Tewolde S, Deyessa N. Effectiveness of a culturally appropriate intervention to prevent intimate partner violence and HIV transmission among men, women, and couples in rural Ethiopia: Findings from a cluster-randomized controlled trial. PLoS Med 2020; 17(8): e1003274.

