## Supplement for "The effect of intimate partner violence on women’s risk of HIV acquisition and engagement in the HIV treatment and care cascade: an individual-participant data meta-analysis of nationally representative surveys in sub-Saharan Africa"

#### Supplement 1: Operational definitions of intimate partner violence and the outcomes of interest

**Table S1.** Operational definitions of physical and/or sexual intimate partner violence (IPV) and indicators most frequently used in surveys included in this analysis.

| | Questions used to define the measures in DHS | Questions used to define the measures in PHIA | Questions used to define the measures in SABSSM |
|---|---|---|---|
| **Lifetime physical IPV** | Did your (last) (husband/partner) ever do any of the following things to you:[§, ¥]<br>● Push you, shake you, or throw something at you?<br>● Slap you?<br>● Punch you with his fist or with something that could hurt you?<br>● Kick you, drag you, or beat you up?<br>● Choke you or burn you on purpose?<br>● Threaten or attack you with a knife, gun, or other weapon? | Not collected | Did your partner ever do any of the following things to you that could hurt you?[§]<br>● Push you, shake you, or throw something at you?<br>● Slap you?<br>● Punch you with his fist or with something<br>● Kick you, drag you, or beat you up?<br>● Try to choke you or burn you on purpose?<br>● Threaten or attack you with a knife, gun, or other weapon? |
| **Lifetime sexual IPV** | Did your (last) (husband/partner) ever do any of the following things to you:<br>● Physically force you to have sexual intercourse with him when you did not want to?<br>● Physically force you to perform any other sexual acts you did not want to?<br>● Force you with threats or in any other way to perform sexual acts you did not want to? | Not collected | Did your partner ever do any of the following things to you that could hurt you?<br>● Physically force you to have sexual intercourse with him/her when you did not want to<br>● Physically force you to perform any other sexual acts you did not want to<br>● Force you with threats or in any other way |
| **Lifetime physical and/or sexual IPV** | All variables in lifetime physical *and* lifetime sexual violence categories | Not collected | All variables in lifetime physical *and* lifetime sexual violence categories |
| **Severe physical and/or sexual IPV** | Did your (last) (husband/partner) ever do any of the following things to you:[§, ¥]<br>● Punch you with his fist or with something that could hurt you?<br>● Kick you, drag you, or beat you up?<br>● Choke you or burn you on purpose?<br>● Threaten or attack you with a knife, gun, or other weapon?<br>Did your (last) (husband/partner) ever do any of the following things to you: | Not collected [#] | Did your partner ever do any of the following things to you that could hurt you? [§]<br>● Punch you with his fist or with something?<br>● Kick you, drag you, or beat you up?<br>● Try to choke you or burn you on purpose?<br>● Threaten or attack you with a knife, gun, or other weapon? |

| | | | |
|---|---|---|---|
| | • Physically force you to have sexual intercourse with him when you did not want to?<br>• Physically force you to perform any other sexual acts you did not want to?<br>• Force you with threats or in any other way to perform sexual acts you did not want to? | | Did your partner ever do any of the following things to you that could hurt you?<br>• Physically force you to have sexual intercourse with him/her when you did not want to?<br>• Physically force you to perform any other sexual acts you did not want to<br>• Force you with threats or in any other way? |
| **Past year physical and/or sexual IPV** | How often did this happen during the last 12 months: often, only sometimes, or not at all?†<br>*Physical violence*<br>• Push you, shake you, or throw something at you?<br>• Slap you?<br>• Punch you with his fist or with something that could hurt you?<br>• Kick you, drag you, or beat you up?<br>• Choke you or burn you on purpose ?<br>• Threaten or attack you with a knife, gun, or other weapon?<br><br>*Sexual violence*<br>• Physically force you to have sexual intercourse with him when you did not want to?<br>• Physically force you to perform any other sexual acts you did not want to?<br>• Force you with threats or in any other way to perform sexual acts you did not want to? | In the past 12 months, did a partner do any of these things to you?<br>*Physical violence*<br>• Slapped you, threw something at you that could hurt you, pushed you or shoved you?<br>• Punched, kicked, whipped, or beat you with an object?<br>• Choked smothered, tried to drown you, or burned you intentionally?<br>• Used or threatened you with a knife, gun or other weapon?<br>*Sexual violence*<br>• In the past 12 months, did a partner physically force you to have sex?<br>• In the past 12 months, did a partner pressure you to have sex and did succeed? | In the last 12 months, how often has your partner *physically* hurt you? ¶ |
| **Frequency of past year physical and/or sexual IPV** | How often did this happen during the last 12 months: often, only sometimes, or not at all?†<br>*Physical violence*<br>• Push you, shake you, or throw something at you?<br>• Slap you?<br>• Punch you with his fist or with something that could hurt you?<br>• Kick you, drag you, or beat you up?<br>• Choke you or burn you on purpose ?<br>• Threaten or attack you with a knife, gun, or other weapon?<br><br>*Sexual violence*<br>• Physically force you to have sexual intercourse with him when you did not want to?<br>• Physically force you to perform any other sexual acts you did not want to?<br>• Force you with threats or in any other way | In the past 12 months, how many times did *someone* ‡<br>• Slapped you, threw something at you that could hurt you, pushed you or shoved you?<br>• Punched, kicked, whipped, or beat you with an object?<br>• Choked smothered, tried to drown you, or burned you intentionally?<br>• Used or threatened you with a knife, gun or other weapon? | In the last 12 months, how often has your partner *physically* hurt you? ¶ |

DHS=Demographic and Health Survey; IPV=intimate partner violence; PHIA=Population-based HIV Impact Assessment Survey; SABSSM=South African National HIV Prevalence, Incidence, Behaviour and Communication Survey.

§ Question “(Does/did) your (last) husband/partner ever twist your arm or pull your hair?” was removed from the definition of lifetime physical IPV to align the DHS, PHIA and SABSSM definitions
¥ In $N_{surv}$= 9 “threatening to attack” and “attack” questions were asked as two separate questions
† In $N_{surv}$ = 13 frequency of past-year IPV was a continuous variable and had to be categorized
‡ In PHIA surveys, the frequency of past-year IPV pertains to only recent physical, not sexual IPV. Furthermore, we assume that “someone” as a perpetrator of violence is an intimate partner only when the woman also reports experiencing IPV.
¶ In SABSSM experience of past year violence, and its frequency pertains to only recent physical, not sexual IPV.
### In PHIA surveys the experience of physical IPV was asked as a single question inquiring about the experience of any of the listed violent acts. This made it impossible to disentangle which women were subjected to severe acts of IPV and which were not.

**Table S2.** Operational definitions and measures of the outcomes used in the analyses.

| Outcome | Numerator | Denominator | Measure |
|---|---|---|---|
| Recent HIV infection | No. recent (past 4-5 months) HIV infection | No. *women who are at risk of HIV infection*, have been randomly selected for IPV module and have ever been in a partnership | Algorithm including limiting Antigen Enzyme (LAg-Avidity) Immunoassay, biomarkers for ART in blood, viral load |
| Lifetime/past year HIV testing | No. tested for HIV ever/in the past 12 months and received the test results | No. women who have been randomly selected for IPV module and have ever been in a partnership | Self-reported |
| ART uptake | No. self-reported current ART intake OR were identified as having ART in blood | No. *HIV positive women* who have been randomly selected for IPV module and have ever been in a partnership | High-resolution liquid chromatography & tandem mass spectrometry |
| Viral load suppression | No. with HIV RNA/mL <1000 copies/mL | No. *HIV positive women* who have been randomly selected for IPV module and have ever been in a partnership | Viral load testing using RT-PCR |

ART=antiretroviral treatment; IPV=intimate partner violence; No.=Number of; RNA=ribonucleic acid; RT-PCR=reverse transcription-polymerase chain reaction.

#### Supplement 2. Missing data analysis for past year physical and/or sexual intimate partner violence and recent HIV infection

##### Missing data on intimate partner violence and recent HIV infections

We calculated missing observations for past year physical and/or sexual intimate partner violence (IPV) and recent HIV infection, as a percentage of the total sample of eligible women for the HIV recency analysis (Table S3). Overall, missing data on IPV was concentrated in one survey – Eswatini 2016 (43% missing). For recent HIV infection, the Uganda 2016 survey has the least amount of missing data for HIV recency (2%) and Malawi has the most (15%).

**Table S3.** Survey-specific frequencies of missing observations for past year physical and/or sexual IPV and recent HIV infection as a proportion of all eligible women for the HIV recency analysis which includes women at risk of HIV acquisition (i.e., excluding those living with non-recent HIV infections).

| | Exposure: Past year physical and/or sexual IPV | | | Outcome: Recent HIV infection | | |
|---|---|---|---|---|---|---|
| **Survey** | **Missing (N)** | **Non missing (N)** | **Proportion missing (%)** | **Missing (N)** | **Non missing (N)** | **Proportion missing (%)** |
| **Overall** | 728 | 22,515 | 3% | 2,651 | 20,582 | 11% |
| Mozambique 2015 | 10 | 2,782 | 0% | 314 | 2,478 | 11% |
| Malawi 2015 | 37 | 6,308 | 1% | 958 | 5,387 | 15% |
| Zambia 2016 | 66 | 5,552 | 1% | 655 | 4,963 | 12% |
| Uganda 2016 | 10 | 1,119 | 1% | 17 | 1,112 | 2% |
| Eswatini 2016 | 560 | 747 | 43% | 96 | 1,211 | 7% |
| Zimbabwe 2015 | 35 | 6,007 | 1% | 611 | 5,431 | 10% |

IPV=intimate partner violence.

##### Justification for a complete case analysis

*Rationale for the proposed missing data mechanism: not missing at random (NMAR)*

Multiple imputation of exposure to past year physical and/or sexual IPV is not warranted for two main reasons. First, if data for past year IPV are not missing at random (NMAR), multiple imputation by chained equations (MICE) would introduce bias in the analysis since MICE assumes missing at random (MAR) or missing completely at random (MCAR).[1] While it is not possible to distinguish between MAR and MNAR based on the available data, evidence has shown that women who have experienced IPV would be less likely to answer survey questions, compared to women not experiencing IPV.[2,3] This could be because women might fear violence by their abuser, should he find out about the interview.[2,3] Thus, missingness is driven by the covariate with the missing data itself, indicating NMAR. Second, the fact that the bulk of the missing data are concentrated in one survey –Eswatini 2016– indicates that a structural, unmeasured factor might have influenced the way that data were collected or recorded, further supporting the argument that data are MAR or MCAR. Still, the amount of missing data does not suggest that IPV module implementation was flawed at large (which was the case for Cameroon PHIA 2017 and Côte d'Ivoire PHIA 2017 surveys which both had over 95% missing data on past year experience of physical and/or sexual IPV), thus warranting inclusion of this survey as part of the analysis. Complete cases analysis is the best option when data are NMAR, unless missingness is outcome driven.[1]

*Estimation of whether missingness of past year IPV is outcome (HIV recency) driven*

To understand if missingness in the exposure is outcome driven, we first compare how the proportion of women with a recent HIV infection differs among those with and without IPV data, and how proportion of women experiencing past year physical and/or sexual IPV differs among women with and without HIV recency data (Table S4). Simple tabulations in Table S4 show that 0.2% of women with both missing and non-missing IPV data had a recent HIV infection.

To investigate the relationship between recent HIV infection and missingness of past year IPV, we ran a logistic regression model with a dummy variable for IPV missingness (0 for missing; 1 for non-missing) as a dependent variable and recent HIV infection as an independent variable. We included survey dummy in the estimation to account for the survey-specific characteristics that might have influenced missingness patterns. This analysis estimated an adjusted odds ratio of 0.73 (95%CI: 0.07-7.14). This effect size estimate is reasonably close to one and, considering the very wide uncertainty, a complete case analysis is warranted.

*Impact of missing data on the estimated relationship between IPV and recent HIV infection*

We investigate exposure to past year IPV among those with missing and non-missing recent HIV infection data (Table S4). 4.5% of women with missing HIV recency data have experienced past year IPV. This proportion is 5.8% among women with non-missing HIV recency data indicating that past year IPV rates are comparable, but slightly higher among women with non-missing HIV recency data. Because we do not know recent HIV infection rates among women with missing HIV recency data, bias could go toward or away from the null.

**Table S4.** Comparison of the proportions of women with a recent HIV infection among those with and without past year physical and/or sexual IPV data; comparison of the proportion of women experiencing past year IPV among those with and without recent HIV infection data.

| **Recent HIV infection** | | **Past year physical and/or sexual IPV** | |
|---|---|---|---|
| **Non-missing IPV, n(%)** | **Missing IPV, n(%)** | **Non-missing recent HIV infection, n(%)** | **Missing recent HIV infection, n(%)** |
| 45/19,935 (0.2%) | 1/647 (0.2%) | 1,158/19,935 (5.8%) | 115/2,580 (4.5%) |

IPV=intimate partner violence.

To investigate the potential relationship between missing HIV recency data and HIV risk, we run a number of regression analyses of various HIV risk factors as dependent variables and non-missing HIV recency status (0 for missing; 1 for non-missing) as independent variables.[4] All models included survey country and year (survey identifier) as a dummy variable to account for survey-specific missingness patterns as shown in previous research.[4] (Table S5)

Women with non-missing HIV recency data were more likely to have earlier sexual debut, had slightly more sexual partners, and were more likely to be currently married (Table S5). Other variables were not associated with the presence of HIV data. Lifetime sexual partners, an earlier sexual debut, and being married might increase the risk of HIV acquisition. Therefore, we suspect that recent HIV infections might be lower among those without HIV recency data.

Because past year physical and/or sexual IPV rates are slightly lower among those without recent HIV data than those with non-missing HIV data (Table S4), the estimated relationship between past year IPV and recent HIV infection could be biased downward (i.e., we underestimate the strength of the effect size). However, we state this with caution given that measurement error and unmeasured confounding could also be influencing the direction of bias.

**Table S5.** Association between non-missing recent HIV infection status and each of the HIV risk factors.

| HIV risk factor | Model type | Estimate | 95%CI |
|---|---|---|---|
| Age at sexual debut | OLS | -0.17 | (-0.29; -0.05) |
| Age | OLS | 0.07 | (-0.39; 0.53) |
| Lifetime number of sexual partners | OLS | 0.01 | (0.01; 0.19) |
| Current marriage status | Logit | 1.03 | (1.01; 1.04) |
| Mean age difference between couple (male age – female age) * | OLS | -0.02 | (-0.36; 0.33) |

95%CI= 95% confidence interval; OLS=Ordinary least squares regression.

*This analysis was conducted using the linked data for married or cohabiting men and women who both declared to be living with each other

#### Supplement 3: Survey-specific burden of intimate partner violence

**Table S6.** Distribution of lifetime physical, lifetime sexual, lifetime physical and/or sexual and past year physical and/or sexual intimate partner violence (IPV) stratified by survey and region

| Country | Survey year | Total sample size | Lifetime physical IPV, N (%) | Lifetime sexual IPV, N(%) | Lifetime physical and/or sexual IPV, N (%) | Past year physical and/or sexual IPV, N (%) |
|---|---|---|---|---|---|---|
| **Overall** | | 280,259 | 73,811 (26.3) | 28,499 (10.2) | 81,555 (29.1) | 59,456 (21.2%) |
| **Central Africa** | | | | | | |
| Angola | 2015 | 7,669 | 2,466 (32.2) | 601 (7.8) | 2,566 (33.5) | 1,886 (24.6) |
| Cameroon | 2004 | 2,184 | 842 (38.6) | 311 (14.2) | 927 (42.4) | 566 (25.9) |
| Cameroon | 2018 | 4,690 | 1,543 (32.9) | 478 (10.2) | 1,643 (35) | 1,001 (21.3) |
| Chad | 2014 | 3,812 | 860 (22.6) | 338 (8.9) | 938 (24.6) | 566 (14.8) |
| Congo, Democratic Republic | 2007 | 2,853 | 1,510 (52.9) | 866 (30.4) | 1,710 (59.9) | 1,589 (55.7) |
| Congo, Democratic Republic | 2013 | 5,689 | 2,564 (45.1) | 1,448 (25.5) | 2,860 (50.3) | 2,126 (37.4) |
| Gabon | 2012 | 4,133 | 1,933 (46.8) | 639 (15.5) | 2,031 (49.1) | 1,315 (31.8) |
| Sao Tome and Principe | 2008 | 1,729 | 501 (29) | 122 (7.1) | 512 (29.6) | 503 (29.1) |
| **Total** | | | **12,219 (37)** | **4,803 (14.6)** | **13,187 (40)** | **9,552 (29.2)** |
| **Western Africa** | | | | | | |
| Burkina Faso | 2010 | 10,003 | 1,125 (11.2) | 150 (1.5) | 1,162 (11.6) | 933 (9.3) |
| Côte d'Ivoire | 2012 | 5,006 | 1,243 (24.8) | 253 (5.1) | 1,279 (25.5) | 1,084 (21.7) |
| Gambia | 2013 | 3,538 | 772 (21.8) | 103 (2.9) | 797 (22.5) | 296 (8.4) |
| Gambia | 2019 | 1,953 | 576 (29.5) | 120 (6.1) | 615 (31.5) | 213 (10.9) |
| Ghana | 2008 | 1,835 | 382 (20.8) | 122 (6.6) | 425 (23.2) | 361 (19.7) |
| Liberia | 2007 | 3,839 | 1,402 (36.5) | 426 (11.1) | 1,516 (39.5) | 1,370 (35.7) |
| Liberia | 2019 | 2,331 | 1,041 (44.7) | 175 (7.5) | 1,064 (45.6) | 820 (35.2) |
| Mali | 2006 | 8,922 | 1,580 (17.7) | 326 (3.7) | 1,676 (18.8) | 1,495 (16.8) |
| Mali | 2012 | 3,120 | 897 (28.7) | 454 (14.6) | 1,069 (34.3) | 816 (26.2) |
| Mali | 2018 | 3,356 | 1,062 (31.6) | 341 (10.2) | 1,128 (33.6) | 638 (19) |
| Nigeria | 2008 | 18,757 | 3,270 (17.4) | 764 (4.1) | 3,431 (18.3) | 2,856 (15.2) |
| Nigeria | 2013 | 22,279 | 3,347 (15) | 1,190 (5.3) | 3,800 (17.1) | 2,600 (11.7) |
| Senegal | 2017 | 2,660 | 529 (19.9) | 173 (6.5) | 599 (22.5) | 344 (12.9) |
| Sierra Leone | 2013 | 4,309 | 1,826 (42.4) | 286 (6.6) | 1,865 (43.3) | 1,201 (27.9) |
| Sierra Leone | 2019 | 4,055 | 1,920 (47.3) | 311 (7.7) | 1,990 (49.1) | 1,490 (36.7) |
| Togo | 2013 | 5,374 | 1,169 (21.8) | 434 (8.1) | 1,271 (23.7) | 737 (13.7) |
| **Total** | | | **22,141 (21.8)** | **5,628 (5.5)** | **23,687 (23.3)** | **17,254 (17)** |
| **Eastern Africa** | | | | | | |
| Burundi | 2016 | 7,366 | 2,826 (38.4) | 1,884 (25.6) | 3,400 (46.2) | 2,088 (28.3) |
| Comoros | 2012 | 2,528 | 136 (5.4) | 43 (1.7) | 156 (6.2) | 119 (4.7) |
| Ethiopia | 2016 | 4,720 | 948 (20.1) | 358 (7.6) | 1,061 (22.5) | 766 (16.2) |
| Kenya | 2003 | 4,312 | 1,662 (38.5) | 606 (14.1) | 1,792 (41.6) | 1,200 (27.8) |
| Kenya | 2008 | 4,901 | 1,767 (36.1) | 716 (14.6) | 1,880 (38.4) | 1,556 (31.7) |
| Kenya | 2014 | 4,515 | 1,590 (35.2) | 529 (11.7) | 1,689 (37.4) | 1,089 (24.1) |
| Malawi | 2004 | 8,292 | 1,672 (20.2) | 1,106 (13.3) | 2,202 (26.6) | 1,591 (19.2) |
| Malawi | 2010 | 3,598 | 776 (21.6) | 616 (17.1) | 1,050 (29.2) | 788 (21.9) |
| Malawi | 2015 | 5,406 | 1,336 (24.7) | 1,020 (18.9) | 1,787 (33.1) | 1,291 (23.9) |
| Mozambique | 2011 | 5,824 | 1,829 (31.4) | 457 (7.8) | 1,926 (33.1) | 1,537 (26.4) |
| Mozambique | 2015 | 3,350 | 622 (18.6) | 133 (4) | 649 (19.4) | 503 (15) |
| Rwanda | 2005 | 2,547 | 786 (30.9) | 356 (14) | 888 (34.9) | 525 (20.6) |

| | | | | | | |
|---|---|---|---|---|---|---|
| Rwanda | 2010 | 3,469 | 1,925 (55.5) | 610 (17.6) | 1,955 (56.4) | 1,576 (45.4) |
| Rwanda | 2015 | 1,907 | 591 (31) | 221 (11.6) | 648 (34) | 395 (20.7) |
| Rwanda | 2019 | 1,947 | 708 (36.4) | 297 (15.3) | 778 (40) | 467 (24) |
| Tanzania | 2010 | 5,689 | 1,917 (33.7) | 731 (12.8) | 2,073 (36.4) | 1,776 (31.2) |
| Uganda | 2006 | 1,748 | 822 (47) | 542 (31) | 985 (56.4) | 752 (43) |
| Uganda | 2011 | 1,702 | 703 (41.3) | 457 (26.9) | 835 (49.1) | 555 (32.6) |
| Uganda | 2016 | 7,536 | 3,088 (41) | 1,710 (22.7) | 3,551 (47.1) | 2,296 (30.5) |
| Zambia | 2007 | 4,230 | 1,853 (43.8) | 742 (17.5) | 2,000 (47.3) | 1,716 (40.6) |
| Zambia | 2013 | 9,412 | 3,628 (38.5) | 1,618 (17.2) | 4,040 (42.9) | 2,589 (27.5) |
| Zambia | 2018 | 7,358 | 2,702 (36.7) | 1,069 (14.5) | 2,962 (40.3) | 1,814 (24.7) |
| Zimbabwe | 2005 | 3,788 | 1,148 (30.3) | 524 (13.8) | 1,333 (35.2) | 1,146 (30.3) |
| Zimbabwe | 2010 | 5,280 | 1,483 (28.1) | 775 (14.7) | 1,795 (34) | 1,383 (26.2) |
| Zimbabwe | 2015 | 5,800 | 1,726 (29.8) | 679 (11.7) | 1,977 (34.1) | 1,137 (19.6) |
| Malawi§ | 2015 | 7,439 | ·· | ·· | ·· | 347 (4.7) |
| Zambia§ | 2016 | 6,591 | ·· | ·· | ·· | 259 (3.9) |
| Uganda§ | 2016 | 1,179 | ·· | ·· | ·· | 136 (11.5) |
| Zimbabwe§ | 2015 | 7,474 | ·· | ·· | ·· | 282 (3.8) |
| **Total** | | | **38,244 (27.3)** | **17,799 (12.7)** | **43,412 (31)** | **31,679 (22.6)** |
| **Southern Africa** | | | | | | |
| Namibia | 2013 | 1,448 | 356 (24.6) | 106 (7.3) | 382 (26.4) | 307 (21.2) |
| South Africa | 2016 | 2,354 | 356 (15.1) | 90 (3.8) | 380 (16.1) | 247 (10.5) |
| Eswatini§ | 2016 | 1,221 | ·· | ·· | ·· | 58 (4.8) |
| South Africa | 2017 | 1,232 | 495 (40.2) | 73 (5.9) | 507 (41.2) | 359 (29.1) |
| **Total** | | | **1,207 (19.3)** | **269 (4.3)** | **1,269 (20.3)** | **971 (15.5)** |

IPV=intimate partner violence.

§ PHIA surveys (5/57) did not collect information on lifetime experience of intimate partner violence (IPV).

#### Supplement 4: Demographic characteristics of women included in each of the analyses for recent HIV infection, HIV testing, antiretroviral uptake, and viral load suppression

**Table S7.** Distribution of demographic characteristics among women included in the analysis for recent HIV infection.

| | Past year physical and/or sexual IPV | |
|---|---|---|
| | **Yes (N=1,273)** | **No (N=21,242)** |
| Recent HIV infection, n (%) | | |
| Yes | 8 (0.6) | 37 (0.2) |
| No | 1,150 (90.3) | 18,740 (88.2) |
| *Missing* | 115 (9) | 2,465 (11.6) |
| Age, n (%) | | |
| 15-24 | 504 (39.6) | 5,612 (26.4) |
| 25-34 | 443 (34.8) | 7,076 (33.3) |
| 35-44 | 188 (14.8) | 4,308 (20.3) |
| 45-64 | 138 (10.8) | 4,246 (20) |
| *Missing* | .. | .. |
| Education, n (%) | | |
| None | 145 (11.4) | 2,516 (11.8) |
| Primary | 699 (54.9) | 11,041 (52) |
| Secondary | 396 (31.1) | 6,626 (31.2) |
| Higher | 30 (2.4) | 873 (4.1) |
| *Missing* | 3 (0.2) | 186 (0.9) |
| Wealth, n (%) | | |
| 1st | 235 (18.5) | 4,529 (21.3) |
| 2nd | 244 (19.2) | 4,295 (20.2) |
| 3rd | 244 (19.2) | 4,098 (19.3) |
| 4th | 255 (20) | 3,951 (18.6) |
| 5th | 295 (23.2) | 4,349 (20.5) |
| *Missing* | .. | 20 (0.1) |
| Residence type, n(%) | | |
| Urban | 496 (39) | 6,603 (31.1) |
| Rural | 777 (61) | 14,639 (68.9) |
| *Missing* | .. | .. |
| Current marital status, n (%) | | |
| Married | 1,027 (80.7) | 17,760 (83.6) |
| Not married | 245 (19.2) | 3,453 (16.3) |
| *Missing* | 1 (0.1) | 29 (0.1) |
| Region, n (%) | | |
| Eastern Africa | 1,247 (98) | 20,521 (96.6) |
| Southern Africa | 26 (2) | 721 (3.4) |
| *Missing* | .. | .. |
| Period, n (%) | | |
| 2015-2019 | 1,273 (100) | 21,242 (100) |
| *Missing* | .. | .. |

IPV=intimate partner violence.

**Table S8.** Distribution of demographic characteristics among women included in the analysis for lifetime and past year HIV testing.

| | Past year physical and/or sexual IPV | |
|---|---|---|
| | **Yes (N=59,456)** | **No (N=217,646)** |
| Lifetime testing for HIV, n(%) | | |
| Yes | 31,023 (52.2) | 113,384 (52.1) |
| No | 28,101 (47.3) | 102,976 (47.3) |
| *Missing* | 332 (0.6) | 1,286 (0.6) |
| Past year HIV testing, n(%) | | |
| Yes | 16,392 (27.6) | 57,996 (26.6) |
| No | 42,601 (71.7) | 157,969 (72.6) |
| *Missing* | 463 (0.8) | 1,681 (0.8) |
| Age, n (%) | | |
| 15-24 | 15,876 (26.7) | 52,458 (24.1) |
| 25-34 | 25,825 (43.4) | 85,515 (39.3) |
| 35-44 | 13,763 (23.1) | 56,359 (25.9) |
| 45-64 | 3,992 (6.7) | 23,314 (10.7) |
| *Missing* | .. | .. |
| Education, n(%) | | |
| None | 17,489 (29.4) | 77,106 (35.4) |
| Primary | 26,901 (45.2) | 78,326 (36) |
| Secondary | 13,896 (23.4) | 52,675 (24.2) |
| Higher | 1,110 (1.9) | 9,156 (4.2) |
| *Missing* | 60 (0.1) | 383 (0.2) |
| Wealth, n(%) | | |
| 1st | 13,966 (23.5) | 47,771 (21.9) |
| 2nd | 12,882 (21.7) | 44,320 (20.4) |
| 3rd | 12,197 (20.5) | 42,453 (19.5) |
| 4th | 11,537 (19.4) | 41,493 (19.1) |
| 5th | 8,515 (14.3) | 40,728 (18.7) |
| *Missing* | 359 (0.6) | 881 (0.4) |
| Residence type, n(%) | | |
| Urban | 18,344 (30.9) | 70,971 (32.6) |
| Rural | 41,112 (69.1) | 146,675 (67.4) |
| *Missing* | .. | .. |
| Current marital status, n(%) | | |
| Married | 53,099 (89.3) | 193,146 (88.7) |
| Not married | 6,355 (10.7) | 24,462 (11.2) |
| *Missing* | 2 (0) | 38 (0) |
| Region, n(%) | | |
| Central Africa | 9,552 (16.1) | 22,622 (10.4) |
| Eastern Africa | 31,679 (53.3) | 106,325 (48.9) |
| Southern Africa | 971 (1.6) | 5,259 (2.4) |
| Western Africa | 17,254 (29) | 83,440 (38.3) |
| *Missing* | .. | .. |
| Period, n(%) | | |
| 2000-2004 | 3,357 (5.6) | 9,871 (4.5) |
| 2005-2009 | 13,869 (23.3) | 40,319 (18.5) |
| 2010-2014 | 23,393 (39.3) | 86,731 (39.8) |
| 2015-2019 | 18,837 (31.7) | 80,725 (37.1) |
| *Missing* | .. | .. |

IPV=intimate partner violence.

**Table S9.** Distribution of demographic characteristics among women included in the analysis for antiretroviral (ART) uptake.

| | Past year physical and/or sexual IPV | |
|---|---|---|
| | **Yes (N=671)** | **No (N=5,278)** |
| ART uptake, n (%) | | |
| Yes | 416 (62) | 3,717 (70.4) |
| No | 232 (34.6) | 1,498 (28.4) |
| *Missing* | 23 (3.4) | 63 (1.2) |
| Age, n (%) | | |
| 15-24 | 92 (13.7) | 479 (9.1) |
| 25-34 | 270 (40.2) | 1,695 (32.1) |
| 35-44 | 202 (30.1) | 1,788 (33.9) |
| 45-64 | 107 (15.9) | 1,316 (24.9) |
| *Missing* | .. | .. |
| Education, n (%) | | |
| None | 26 (3.9) | 448 (8.5) |
| Primary | 209 (31.1) | 2,257 (42.8) |
| Secondary | 364 (54.2) | 2,184 (41.4) |
| Higher | 20 (3) | 207 (3.9) |
| *Missing* | 52 (7.7) | 182 (3.4) |
| Wealth, n (%) | | |
| 1st | 49 (7.3) | 782 (14.8) |
| 2nd | 36 (5.4) | 735 (13.9) |
| 3rd | 45 (6.7) | 832 (15.8) |
| 4th | 96 (14.3) | 999 (18.9) |
| 5th | 86 (12.8) | 1,069 (20.3) |
| *Missing* | 359 (53.5) | 861 (16.3) |
| Residence type, n (%) | | |
| Urban | 318 (47.4) | 2,245 (42.5) |
| Rural | 353 (52.6) | 3,033 (57.5) |
| *Missing* | .. | .. |
| Current marital status, n (%) | | |
| Married | 292 (43.5) | 2,945 (55.8) |
| Not married | 378 (56.3) | 2,324 (44) |
| *Missing* | 1 (0.1) | 9 (0.2) |
| Region, n (%) | | |
| Eastern Africa | 280 (41.7) | 3,979 (75.4) |
| Southern Africa | 391 (58.3) | 1,299 (24.6) |
| *Missing* | .. | .. |
| Period, n (%) | | |
| 2015-2019 | 671 (100) | 5,278 (100) |
| *Missing* | .. | .. |

ART=antiretroviral treatment; IPV=intimate partner violence.

**Table S10.** Distribution of demographic characteristics among women included in the analysis for viral load suppression (VLS).

| | **Past year physical and/or sexual IPV** | |
|---|---|---|
| | **Yes (N=671)** | **No (N=5,278)** |
| Viral load suppression, n (%) | | |
| Yes | 375 (55.9) | 3,506 (66.4) |
| No | 286 (42.6) | 1,700 (32.2) |
| *Missing* | 10 (1.5) | 72 (1.4) |
| Age, n (%) | | |
| 15-24 | 92 (13.7) | 479 (9.1) |
| 25-34 | 270 (40.2) | 1,695 (32.1) |
| 35-44 | 202 (30.1) | 1,788 (33.9) |
| 45-64 | 107 (15.9) | 1,316 (24.9) |
| *Missing* | .. | .. |
| Education, n (%) | | |
| None | 26 (3.9) | 448 (8.5) |
| Primary | 209 (31.1) | 2,257 (42.8) |
| Secondary | 364 (54.2) | 2,184 (41.4) |
| Higher | 20 (3) | 207 (3.9) |
| *Missing* | 52 (7.7) | 182 (3.4) |
| Wealth, n (%) | | |
| 1st | 49 (7.3) | 782 (14.8) |
| 2nd | 36 (5.4) | 735 (13.9) |
| 3rd | 45 (6.7) | 832 (15.8) |
| 4th | 96 (14.3) | 999 (18.9) |
| 5th | 86 (12.8) | 1,069 (20.3) |
| *Missing* | 359 (53.5) | 861 (16.3) |
| Residence type, n (%) | | |
| Urban | 318 (47.4) | 2,245 (42.5) |
| Rural | 353 (52.6) | 3,033 (57.5) |
| *Missing* | .. | .. |
| Current marital status, n (%) | | |
| Married | 292 (43.5) | 2,945 (55.8) |
| Not married | 378 (56.3) | 2,324 (44) |
| *Missing* | 1 (0.1) | 9 (0.2) |
| Region, n (%) | | |
| Eastern Africa | 280 (41.7) | 3,979 (75.4) |
| Southern Africa | 391 (58.3) | 1,299 (24.6) |
| *Missing* | .. | .. |
| Period, n (%) | | |
| 2015-2019 | 671 (100) | 5,278 (100) |
| *Missing* | .. | .. |

IPV=intimate partner violence; VLS=viral load suppression.

**Supplement 5: Analysis of couple and co-habiting male partner characteristics**

We performed robustness checks and examined the characteristics of the women's cohabiting partner as potential confounders of the association in the six surveys included in the recent HIV infection analysis.

*Partners' HIV status*

Despite the small resulting sample size, (only two women were both subjected to past year IPV and had a recent HIV infection), the HIV prevalence among cohabiting partners of women who had experienced past year IPV was similar (16%) to that of partners of women who had not (14%) (Table S11). Similarly, proportions of condom use at women's last sex was similar and low between women subjected to (11%) and not subjected to (10%) recent IPV (Table S12). Distribution of male partner education was also comparable between women subjected to and not subjected to recent IPV.

*Partners' educational levels*

There were no large differences in the partner's education level and IPV. Majority of the male partners of women experiencing IPV in the past year (51.8%), as well as those not experiencing IPV (47.8%) had primary education.

*Couple age discrepancy*

Partner age discrepancy was slightly larger among women who had not reported past year IPV compared to those who had, though the difference was only 0.6 years. We adjusted for couple age discrepancy in a sensitivity analysis. This did not change the effect estimate though the precision was reduced since the sample size was halved (aPR=3·24, 95%CI: 0·72-14·63, $N_{surv}$=6; Table S11). Furthermore, this analysis includes only currently cohabiting women who mutually declare to be living with their partner.

*Partners' alcohol consumption*

Another potential confounder of the relationship between IPV and HIV acquisition is the male partner alcohol consumption (Table S11). We found that the proportion of men drinking alcohol "Often" is higher among women experiencing past year IPV (20%) compared to women not experiencing IPV (15%). Similarly, the proportion of men "Never" drinking alcohol is higher among women not subjected to past year IPV (54%) compared to women subjected to IPV (38%). These results are aligned with previous meta-analyses demonstrating that alcohol and IPV perpetration are often associated.[5,6] If there is an association between male alcohol consumption and HIV acquisition among women, this variable might be confounding the IPV-HIV incidence relationship.

We explore this further by examining the distribution of recent HIV infections by male partner's frequency of alcohol consumption. Women whose partners "Often" drink have higher proportion of incident HIV (0.11%) compared to women whose partner "Rarely" drinks (0.06%) (Table S13). Question remains on pathways through which alcohol consumption effects HIV acquisition. Among the 12 women with incident HIV, half of their male partners ($N_i$=6) are HIV positive and virally unsuppressed. Alcohol use can lead to poor ART adherence and subsequently, poor viral load suppression.[7] In our sample of HIV positive male partners, men who drink "Often" are more likely to be virally unsuppressed (51.3%) compared to other frequency categories. Men who "Never" drink are more likely to be virally suppressed (63.7%) (Table S14).

If male partner's alcohol consumption leads to women HIV's acquisition through poor male ART adherence and viral suppression, it would be sufficient to control for this latter variable to obtain an adjusted effect size estimate for IPV (Figure S1). In addition, we have more surveys with information on viral load suppression and this variable is objectively measured via biomarkers. In comparison, questions of alcohol consumptions are self-reported and for them the questions are not standardized across surveys. After adjusting for male partner viral load suppression in the subset of HIV positive men ($N_i$=1,505) the effect estimate remains robust though the CI is large due to a reduction in the sample size by 92% (aPR = 4.87 95%CI: (0.81-29.45)).

**Table S11.** Unweighted proportions of couple and male partner characteristics stratified by experience of past year intimate partner violence (IPV), among the six surveys (Mozambique, Malawi, Zambia, Uganda, Eswatini, Zimbabwe) containing information on a recent HIV infection and cohabiting partners.

| | **Experiencing past year physical and/or sexual IPV** | **Not experiencing past year physical and/or sexual IPV** |
|---|---|---|
| Male partner HIV status, n (%)§ | | |
| Positive | 112 (16.2) | 1,544 (13.6) |
| Negative | 548 (79.3) | 8,735 (77.2) |
| *Missing* | 31 (4.5) | 1,035 (9.1) |
| Male partner education, n(%) | | |
| None | 30 (4.3) | 612 (5.4) |
| Primary | 358 (51.8) | 5,408 (47.8) |
| Secondary | 256 (37.0) | 4,380 (38.7) |
| Higher | 42 (6.1) | 795 (7.0) |
| *Missing* | 5 (0.7) | 119 (1.1) |
| Couple age discrepancy (man - woman) stratified by women's age, mean (SD)¥ | | |
| 15-24 | 6.21 (5.62) | 6.25 (4.88) |
| 25-34 | 5.58 (5.07) | 6.15 (5.26) |
| 35-44 | 4.91 (5.99) | 6.15 (6.04) |
| 45-64 | 2.98 (5.64) | 6.02 (6.28) |
| Male partner alcohol consumption, n(%)‡ | | |
| Never | 259 (37.5) | 6,145 (54.3) |
| Sometimes | 204 (29.5) | 2,845 (25.1) |
| Often | 139 (20.1) | 1,695 (15.0) |
| *Missing* | 89 (12.9) | 629 (5.6) |

SD=standard deviation; IPV=intimate partner violence.

§ Difference in partner HIV status between women experiencing and not experiencing past year intimate partner violence (IPV) was not statistically significant ($p<0.05$)

‡ Question on the frequency of alcohol consumption was not asked in Uganda 2016 PHIA survey. In PHIA surveys the question about alcohol consumption was asked to the male partner directly; in Mozambique 2015 AIS survey, women were asked about their partners' alcohol consumption frequency.

¥ Difference in the mean couple age discrepancy between women who had not experienced past year physical and/or sexual IPV and women who had was less than one year (0.6 years).

**Table S12.** Unweighted proportion of women who used condoms during last sex in the last 12 months, stratified by experience of past year physical and/or sexual intimate partner violence (IPV), among the six surveys (Mozambique, Malawi, Zambia, Uganda, Eswatini, Zimbabwe) with available information on condom use during the woman's last sexual act.

| | **Experiencing past year physical and/or sexual IPV** | **Not experiencing past year physical and/or sexual IPV** |
|---|---|---|
| Condom use during woman's most recent sex, n (%) | | |
| Use | 77 (11.1) | 1,172 (10.4) |
| No use | 578 (83.6) | 9,166 (81.0) |
| *Missing* | 36 (5.2) | 976 (8.6) |

IPV=intimate partner violence.

**Table S13.** Unweighted proportion of recent HIV infection among women, stratified by male partner alcohol consumption frequency among the six surveys (Mozambique, Malawi, Zambia, Uganda, Eswatini, Zimbabwe) with available information.

| | **Male partner's frequency of alcohol consumption**‡ | | |
|---|---|---|---|
| | **Never** | **Sometimes** | **Often** |
| Recent HIV infection, n (%)§ | | | |
| Recent HIV infection | 4 (0.06) | 5 (0.16) | 2 (0.11) |
| Non-recent HIV infection | 962 (14.3) | 489 (15.5) | 260 (13.9) |
| Not living with HIV | 5,207 (77.5) | 2,434 (77.3) | 1,487 (79.4) |
| *Missing* | 543 (8.1) | 221 (7.0) | 123 (6.6) |

§ One woman with incident HIV is removed from the analysis because she had a missing value for male partner alcohol consumption

‡ The denominators in this analysis are the male partners of women (regardless of men's HIV status) who have declared to be in a cohabitating partnership and are at risk of HIV acquisition.

**Table S14.** Unweighted proportion of HIV positive male partners who are virally suppressed stratified by frequency of their alcohol consumption among the six surveys (Mozambique, Malawi, Zambia, Uganda, Eswatini, Zimbabwe) with available information.

| | **Male partner's frequency of alcohol consumption**‡ | | |
|---|---|---|---|
| | **Never** | **Sometimes** | **Often** |
| Viral load suppression, n (%) | | | |
| Suppressed | 654 (63.7) | 293 (59.0) | 113 (47.9) |
| Unsuppressed | 365 (35.6) | 196 (39.4) | 121 (51.3) |
| *Missing* | 7 (0.7) | 8 (1.6) | 2 (0.8) |

IPV=intimate partner violence.

‡ The denominators in this analysis are *HIV positive* male partners of women who have declared to be in a cohabitating partnership and are at risk of HIV acquisition.

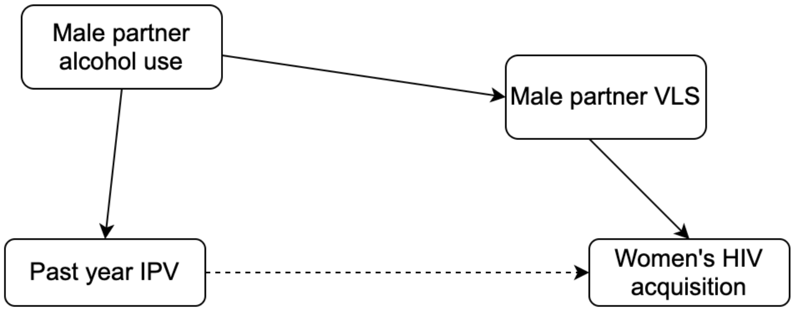

**Figure S1**: Directed Acyclic Graph (DAG) of the proposed relationship between past year IPV and women's HIV acquisition.

(DAG= directed acyclic graph; IPV=intimate partner violence; VLS=viral load suppression)

#### Supplement 6: Sensitivity analyses

**Table S15.** Crude and adjusted prevalence ratios of lifetime and past year HIV testing among women delivering outside the antenatal care (ANC) and experiencing different intimate partner violence (IPV) categories compared to women not experiencing a specific type of IPV.

| **Outcome** | **Exposures** | **$N_{surv}$** | **$N_i$** | **Crude PR (95% CI)** | **Adjusted PR§ (95% CI)** |
|---|---|---|---|---|---|
| ***Past year HIV testing*** | | | | | |
| **Primary exposure of interest** | | | | | |
| | Past year physical and/or sexual IPV | 57 | 198,979 | 0.97 (0.95, 0.99) | 1.00 (0.98, 1.03) |
| **Secondary exposure of interest** | | | | | |
| | Lifetime physical IPV | 52 | 188,798 | 1.03 (1.01, 1.05) | 1.01 (0.99, 1.03) |
| | Lifetime sexual IPV | 52 | 188,769 | 1.05 (1.02, 1.07) | 1.00 (0.97, 1.02) |
| | Lifetime physical and/or sexual IPV | 52 | 188,775 | 1.04 (1.02, 1.06) | 1.01 (0.99, 1.03) |
| | Severe lifetime physical and/or sexual IPV† | 52 | 188,743 | 1.03 (1.01, 1.05) | 1.00 (0.98, 1.02) |
| | Frequency of past-year physical and/or sexual IPV¥ | | | | |
| | Never | 57 | 198,979 | Referent | Referent |
| | Sometimes | | | 0.97 (0.96, 0.99) | 1.00 (0.98, 1.03) |
| | Often | | | 0.96 (0.93, 0.99) | 1.01 (0.97, 1.05) |
| ***Lifetime HIV testing*** | | | | | |
| **Secondary exposure of interest** | | | | | |
| | Lifetime physical IPV | 52 | 188,958 | 1.03 (1.02, 1.04) | 1.04 (1.03, 1.05) |
| | Lifetime sexual IPV | 52 | 188,928 | 1.03 (1.02, 1.04) | 1.01 (1.00, 1.03) |
| | Lifetime physical and/or sexual IPV | 52 | 188,935 | 1.04 (1.03, 1.05) | 1.04 (1.03, 1.05) |
| | Severe lifetime physical and/or sexual violence IPV† | 52 | 188,902 | 1.02 (1.01, 1.03) | 1.02 (1.00, 1.03) |

95%CI=95% confidence intervals; ANC=antenatal care; IPV=intimate partner violence; $N_{surv}$=number of surveys; $N_i$=number of individual observations without any missing data for intimate partner violence (IPV), the outcome and the covariates included in the model; PR=prevalence ratio.

§ Adjusted for: age(continuous), residence type (rural/urban), women's current marital status (married/unmarried), women's education (none, primary, secondary, higher), and survey identifier.

¥ In PHIA surveys (5/57) the frequency of past-year intimate partner violence (IPV) pertains to only recent physical, not sexual IPV. Furthermore, this question asks about perpetration of violence by "someone" which we assume to be an intimate partner only when a woman also reports experiencing IPV. The referent category for the frequency of past year physical and/or sexual IPV are women who did not experience any past year physical and/or sexual intimate partner violence (IPV).

† Referent category includes women experiencing non-severe lifetime IPV and no lifetime IPV.

**Table S16.** Crude and adjusted prevalence ratios of viral suppression among women living with HIV (WLHIV) on antiretroviral treatment (ART) experiencing different intimate partner violence (IPV) categories compared to women not experiencing a specific type of IPV.

| **Exposures** | $N_{surv}$ | $N_i$ | **Crude PR (95% CI)** | **Adjusted PR§ (95% CI)** |
|---|---|---|---|---|
| **Primary exposure of interest** | | | | |
| Past year physical and/or sexual IPV | 7 | 3,932 | 0.91 (0.87, 0.95) | 0.94 (0.9, 0.99) |
| **Secondary exposures of interest** | | | | |
| Lifetime physical IPV | 2 | 942 | 0.98 (0.93, 1.05) | 0.94 (0.89, 1.00) |
| Lifetime sexual IPV | 2 | 943 | 1.03 (0.92, 1.15) | 1.01 (0.90, 1.14) |
| Lifetime physical and/or sexual IPV | 2 | 942 | 0.99 (0.93, 1.05) | 0.95 (0.89, 1.01) |
| Severe lifetime physical and/or sexual IPV† | 2 | 942 | 1.04 (0.96, 1.12) | 1.03 (0.95, 1.12) |
| Frequency of past-year physical and/or sexual IPV¥ | | | | |
| Never | 7 | 3,932 | Referent | Referent |
| Sometimes | | | 0.93 (0.89, 0.98) | 0.96 (0.91, 1.01) |
| Often | | | 0.77 (0.64, 0.92) | 0.86 (0.72, 1.02) |

95%CI=95% confidence intervals; ART=antiretroviral treatment; IPV=intimate partner violence; $N_{surv}$=number of surveys; $N_i$=number of individual observations without any missing data for intimate partner violence (IPV), the outcome and the covariates included in the model; PR=prevalence ratio; WLHIV=women living with HIV.

§ Adjusted for: age(continuous), residence type (rural/urban), women's current marital status (married/unmarried), women's education (none, primary, secondary, higher), and survey identifier.

¥ In PHIA surveys (5/57) the frequency of past-year intimate partner violence (IPV) pertains to only recent physical, not sexual IPV. Furthermore, this question asks about perpetration of violence by "someone" which we assume to be an intimate partner only when a woman also reports experiencing IPV. The referent category for the frequency of past year physical and/or sexual IPV are women who did not experience any past year physical and/or sexual intimate partner violence (IPV).

† Referent category includes women experiencing non-severe lifetime IPV and no lifetime IPV.
